# Atypical depression is associated with a distinct clinical, neurobiological, treatment response and polygenic risk profile

**DOI:** 10.1101/2025.08.05.25333073

**Authors:** Mirim Shin, Jacob J. Crouse, Tian Lin, Enda M Byrne, Brittany L. Mitchell, Penelope A. Lind, Richard Parker, Sarah Mckenna, Emiliana Tonini, Joanne S. Carpenter, Kathleen R. Merikangas, Naomi R. Wray, Sarah E. Medland, Nicholas G. Martin, Ian B. Hickie

**Affiliations:** Brain and Mind Centre, The University of Sydney, Australia; Institute for Molecular Bioscience, The University of Queensland, Brisbane, QLD, Australia; Child Health Research Centre, The University of Queensland, Brisbane, QLD, Australia; Brain and Mental Health Program, QIMR Berghofer Medical Research Institute, Brisbane, QLD, Australia; School of Biomedical Sciences, Queensland University of Technology, Brisbane, Australia; Faculty of Medicine, School of Biomedical Sciences, University of Queensland, Brisbane, Australia; Genetic Epidemiology Research Branch, Intramural Research Program, National Institute of Mental Health, Bethesda, MD, USA; Department of Psychiatry, University of Oxford, Oxford, United Kingdom; Li Ka Shing Centre for Health Information and Discovery, University of Oxford, Oxford, United Kingdom

**Keywords:** depression, mood disorder, polygenic, immune, immuno-metabolic, circadian

## Abstract

**Background:** Atypical depression is considered a distinct clinical subtype of major depression, yet its predictive validity and clinical utility remain contested. We investigated association between atypical depression and clinical characteristics, genetic profiles, and antidepressant responses.

**Methods:** Among 14,897 participants from the Australian Genetics of Depression Study (75% female; mean age 43.7 years), 3,098 (21%) were classified phenotypically as having “atypical depression” based on self-reported weight gain and hypersomnia during their worst depressive episode. Demographics and clinical features were compared. Bonferroni-corrected regression models assessed associations between atypical depression and polygenic scores (PGS) for mental disorders, metabolic-inflammatory-circadian traits, and self-reported antidepressant response and side effects.

**Results:** Atypical depression cases had an earlier age of onset, greater illness severity, stronger eveningness and reduced daylight exposure. Atypical depression cases had higher PGS for major depressive disorder (odds ratio [OR]=1.10 [95%CI: 1.06-1.15]), attention-deficit/hyperactivity disorder (OR=1.08 [1.04-1.13]), bipolar disorder (OR= 1.07 [1.02-1.12]), neuroticism (OR=1.07 [1.02-1.12]), BMI (OR=1.35 [1.29-1.42]), Type 2 diabetes (OR=1.22 [1.16-1.28]), C-reactive protein (OR=1.12 [1.07-1.17]), and insulin resistance (OR=1.11 [1.06-1.16]) but lower PGS for HDL cholesterol (OR=0.91 [0.87-0.95]) and chronotype (indicating eveningness; OR=0.94 [0.90-0.98]). Atypical depression was associated with poorer efficacy of selective serotonin reuptake inhibitors (OR=0.88 [0.81-0.96]) and serotonin-norepinephrine reuptake inhibitors (OR=0.85 [0.77-0.94]), along with more side effects, particularly weight gain (OR=2.89 [2.66-3.15]).

**Conclusions:** This large genetically-informative study supports the neurobiological and clinical validity of atypical depression, demonstrating distinct clinical and genetic risk profiles alongside differential antidepressant responses. These support utilizing the atypical subtype to guide treatment selection and physical health management.

## INTRODUCTION

The concept of atypical depression has evolved over 60 years,^1^ initially describing patients preferentially responsive to monoamine oxidase inhibitors (MAOIs) while relatively unresponsive to tricyclic antidepressants (TCAs) or electroconvulsive therapy (ECT).^1^ The Diagnostic and Statistical Manual of Mental Disorders (DSM-5-TR) classifies atypical depression by mood reactivity *plus* at least two features: increased appetite/weight gain, hypersomnia, leaden paralysis, and interpersonal rejection sensitivity.^2^ Debate persists regarding the relevance and reliable assessment of mood reactivity^3,4^ and rejection sensitivity.^4–6^ Importantly, the core feature of mood reactivity is poorly differentiating and lacks consistent associations with other atypical features^6–8^ and treatment response.^4,9^ By contrast, the reversed neurovegetative symptoms (i.e., hypersomnia and increased appetite/weight gain) have emerged empirically as more robust markers, showing stronger biological correlates and greater clinical significance.^10,11^ These features are particularly prominent in severe, recurrent depression,^12,13^ and are widely used in epidemiologic studies.^14–16^

An important measure of the utility of the atypical subtype is whether it can inform treatment selection. Currently the evidence of MAOI superiority over other antidepressants in atypical depression is mixed. Some studies show comparable efficacy between serotonin reuptake inhibitors (SSRIs) (e.g., fluoxetine, sertraline) and MAOIs (e.g., phenelzine, moclobemide)^17,18^ whereas others report poorer SSRIs response.^19,20^ Meta-analyses have reported MAOI superiority over TCAs with a medium effect size, but MAOI advantage over SSRIs in atypical depression remains unclear.^21,22^

Atypical depression is associated clinically with higher female prevalence, earlier onset, greater illness severity, higher recurrence, and more significant impairment.^5,15,23–26^ Biologically, it associates with decreased hypothalamic-pituitary-adrenal axis activity,^4,27,28^ elevated immune mediators,^29,30^ and metabolic dysregulation, leading to higher diabetes and cardiovascular disease risks.^31–36^ In youth, these changes often precede weight gain,^37^ and include increased inflammatory markers such as C-reactive protein (CRP), interleukin-6 (IL-6), and tumor necrosis factor-α (TNF-α).^38,39^ Some studies suggest a neurobiological link with circadian factors, however this is underexplored.^40,41^

Genetic studies reinforce these distinctions, linking atypical depression to higher polygenic scores (PGS) for metabolic-inflammatory traits, particularly for CRP, BMI, leptin, and triglycerides.^10,11,36,42^ However, broader immune-inflammatory-related PGS remain underexplored.^42^ Despite higher comorbid mental disorder rates,^12^ the association between PGS for various mental disorders and atypical depression remains mixed.^10,36,42,43^ While ‘non-atypical’ depression shows stronger genetic overlap with schizophrenia,^10,44^ atypical depression is more strongly associated with bipolar disorders^14^ and ADHD,^36^ suggesting distinct genetic risk profiles.

To address the ongoing debate about atypical depression’s neurobiological and clinical validity, this study leveraged one of the world’s largest genetically informative depressive disorder cohorts (Australian Genetics of Depression Study [AGDS]).^45^ Using reversed neurovegetative symptoms (increased weight gain and hypersomnia) as the defining criteria, we hypothesized that adults with atypical depression, compared to adults with other depressive disorders, will show:

1. different demographic and clinical characteristics (e.g., higher female proportion, more severe illness with comorbidity, earlier onset, delayed sleep-wake schedules, evening chronotype);
2. higher PGS for some mental health traits or disorders (e.g., Major Depressive Disorder [MDD], Attention-Deficit Hyperactivity Disorder [ADHD], bipolar disorder and neuroticism) but not all (e.g., Schizophrenia);
3. higher PGS for metabolic markers (e.g., BMI, fasting insulin, Type 2 Diabetes [T2D], High-Density Lipoprotein Cholesterol [HDL-C], Insulin resistance ([IR] derived two ways: from the Tryglycedirde:HDL-C ratio and from glucose-insulin indices [HOMA-IR]) and Coronary Artery Disease [CAD]), immune-related markers (e.g., CRP, IL-6, and TNF-α);
4. significant association with sleep/circadian markers (e.g., higher sleep midpoint, lower chrontype, etc.)
5. lower self-reported efficacy of common antidepressants (SSRIs, Serotonin-Norepinephrine Reuptake Inhibitors [SNRIs], and TCAs); and
6. higher overall rates of side effects, notably weight gain, across antidepressant classes.

These hypotheses were not pre-registered.

## METHODS AND MATERIALS

### Participants and Study Design

Participants were from the AGDS, a cohort study exploring genetic and psychosocial factors influencing depressive disorder etiology and antidepressant response. A cohort profile is described elsewhere.^45^ Data from the first freeze (September 2016-September 2018) were analyzed. Ethical approval was granted by the QIMR Berghofer Medical Research Institute Human Research Ethics Committee in Brisbane, Australia, with written informed consent obtained. This study followed the Strengthening the Reporting of Observational Studies in Epidemiology (STROBE) guideline.

20,689 participants (75% female; mean age 43±15 years [range: 18-90]) with a self-reported diagnosis of, or treatment for, a depressive disorder were recruited; 76% provided a saliva sample. Over 95% were of genetically inferred European ancestry and PGS were created for European ancestry participants only. Participants completed online surveys comprising a core module on depressive symptomatology and medication experiences, and additional modules, including chronotype and daylight exposure during workdays and free days (see Supplementary Materials).

MDD was operationalized using DSM-5 criteria. The AGDS cohort predominantly comprises severe, recurrent depression cases, with only 4% reporting one depressive episode.^45^ Consistent with other studies that rely on self-report rather than clinically-administered instruments,^12,13^ those reporting both weight gain *and* hypersomnia during their worst depressive episode were classified as having atypical depression, given stronger and more consistent biological marker associations than other atypical features. The core DSM-5 ‘mood reactivity’ criterion was not assessed, as this clinical concept requires direct reporting or observation during depressive episodes, which was not feasible given retrospective, self-report nature of the AGDS study design. Increased appetite was not recorded. All other cases were categorized as “other depressive disorders”.

### Polygenic Scores (PGS)

DNA samples were collected using saliva kits, genotyped using the Illumina Global Screening Array V.2.0. Pre-imputation quality control used PLINK 1.9.^46,47^ Quality control removed SNPs with a minor allele frequency <0.005, SNP call rate <97.5%, and Hardy-Weinberg equilibrium (p<1×10^−6^), before imputation using the TopMed r2 reference panel.^48^ Ancestry was inferred using the first three principal components (PCs), by projecting PCs on 1000 Genomes data using GCTA.^49^ Samples within six standard deviations (SD) of each ancestry’s PC in 1000 Genomes were assigned as the same ancestry. The latest publicly available genome-wide association study (GWAS) results for mental disorders, metabolic, inflammatory, and circadian traits (see Supplementary Materials) were used as weights in the calculation of PGS. Where applicable, leave-one-out summary statistics were used for GWAS studies that included participants from AGDS to avoid over-estimation. SBayesRC was used to generate allele weights for each PGS.^50^ The posterior SNP effects for each disorder/trait were used to generate PGS for each participant using the *PLINK* score function.^46^ Each PGS was standardized using the scale() function. Effect sizes are interpretable as SD units of AGDS PGS. In our previous analysis of this cohort,^51^ SDs were similar between AGDS cases and the control cohort, but mean PGS values (particularly for MDD) were significantly higher in AGDS, consistent with the case-control study design.

### Antidepressants: efficacy and side effects

The survey assessed participants’ experiences with 10 common antidepressants. Efficacy (i.e., how well each antidepressant worked) was rated on an ordinal scale (0=“not at all well”, 1=“moderately well”, 2=“very well”), excluding “I don’t know” responses. Efficacy was compared across three major classes: SSRIs (sertraline, escitalopram, citalopram, fluoxetine, paroxetine), SNRIs (venlafaxine, desvenlafaxine, duloxetine), and TCAs (amitriptyline, mirtazapine). For differing efficacy ratings within the same class, the highest efficacy-rating was used. Side effects were assessed as a binary variable (0=“no”, 1=“yes”) across 25 possible symptoms. Participants reported side effects only for antidepressants they had taken.

### Statistical Analysis

Analyses were conducted in RStudio (R version 4.5.0).^52^ Demographic and clinical characteristics of atypical vs other depressive disorders were compared using Welch’s t-test for age, Analysis of Covariance (adjusting for age and sex) for other continuous variables, and Pearson’s Chi-squared test for categorical variables. For these exploratory comparisons, multiple testing was controlled using the false discovery rate, with adjusted p-values (q<0.05) considered significant. Complete case analysis was used without imputation, which may introduce bias. For PGS analyses, logistic regression estimated associations with atypical depression, adjusting for age, sex, and the first 10 genetic PCs (to account for population stratification). Treatment response analyses used logistic regression for side effects (across 10 antidepressants) and ordinal regression for antidepressant efficacy, adjusting for age and sex. Bonferroni correction was applied to hypothesis-driven analyses, requiring stricter control of Type I error:

1. *Genetic:* Logistic regression tested associations between atypical depression and three PGS sets: (i) mental disorders (Bonferroni-p-threshold: 0.0071 for seven PGS), (ii) sixteen metabolic/inflammatory traits (Bonferroni-p-threshold: 0.0031), and (iii) five sleep/circadian traits (Bonferroni-p-threshold: 0.01).
2. *Antidepressant Efficacy:* Ordinal regression assessed atypical depression’s effect on reported efficacy of three classes of medications (SSRIs, SNRIs, TCAs; Bonferroni-p-threshold: 0.0167 (0.05/3)).
3. *Side effects:* Logistic regression examined associations with 25 side effects (Bonferroni-p-threshold: 0.002 (0.05/25)).

## RESULTS

### Demographic and clinical characteristics

Of 14,897 participants (75% female; 43.7±15.3 years) with available genetic data passing quality control who answered questions about weight and sleep changes during their worst depressive episode, 21% (n=3,098) met our case definition for atypical depression.

Compared to “other depressive disorders”, atypical depression cases were younger (42.9±14.4 vs 43.9±15.6, q=0.002), had higher BMI (31.8±6.7 vs 27.2±6.2, q<0.001; noting 31% missingness), and were more often females (79% vs 73%, q<0.001, Table 1). Among all participants, 88% met DSM-5 criteria for a lifetime major depressive episode (MDE). Clinically, atypical depression cases had earlier onset (21.7±10.7 vs 22.3±11.5, q=0.003), higher MDE rates (99.6% vs 84.8%) and generalized anxiety disorder (53% vs 48%), reported more lifetime depressive episodes, and greater severity across questionnaires for hypo/mania, psychosis, suicidality, psychological distress, eveningness preference, seasonality, and less time spent outdoors on workdays and free days (all q’s<0.001). Atypical cases had significantly higher severe substance use disorder rates (particularly nicotine and drug use), and increased prevalence of diabetes (8% vs 5%) and high blood pressure (18% vs 14%) (all q’s<0.001).

**Table 1.** Demographic and clinical characteristics of the included sample (N=14,897)

| **Characteristic** | **Total sample**<br>**(N=14,897)** | **Non-Atypical**<br>**(n=11,799)** | **Atypical**<br>**(n=3,098)** | **p-value** | **q-value** |
| --- | --- | --- | --- | --- | --- |
| Age (years) | 43.70±15.32 | 43.90±15.55 | 42.93±14.41 | 0.001 | 0.002 |
| Sex |  |  |  | <0.001 | <0.001 |
| Female | 11,088 (74.4) | 8,655 (73.4) | 2,433 (78.5) |  |  |
| Male | 3,809 (25.5) | 3,144 (26.6) | 665 (21.5) |  |  |
| Information not provided | 14 (0.1) | 10 (0.1) | 4 (0.1) |  |  |
| BMI | 28.15±6.53 | 27.19±6.15 | 31.75±6.67 | <0.001 | <0.001 |
| Information not provided | 4,684 (31.4) | 3,739 (31.7) | 945 (30.5) |  |  |
| Marital status |  |  |  | <0.001 | <0.001 |
| Married or de facto relationship | 7,976 (53.5) | 6,457 (54.7) | 1,519 (49.0) |  |  |
| Never married | 4,389 (29.5) | 3,414 (28.9) | 975 (31.5) |  |  |
| Separated or divorced | 2,245 (15.1) | 1,698 (14.4) | 547 (17.7) |  |  |
| Widowed | 255 (1.7) | 208 (1.8) | 47 (1.5) |  |  |
| Information not provided | 32 (0.2) | 22 (0.2) | 10 (0.3) |  |  |
| Education |  |  |  | 0.013 | 0.017 |
| Postgraduate | 4,123 (27.7) | 3,306 (28.0) | 817 (26.4) |  |  |
| Degree | 5,198 (34.9) | 4,144 (35.1) | 1,054 (34.0) |  |  |
| Certificate or diploma | 3,509 (23.6) | 2,713 (23.0) | 796 (25.7) |  |  |
| Senior high school | 1,181 (7.9) | 917 (7.8) | 264 (8.5) |  |  |
| Junior high school or less | 854 (5.7) | 692 (5.9) | 162 (5.2) |  |  |
| No formal education | 7 (0.0) | 6 (0.1) | 1 (0.0) |  |  |
| Information not provided | 25 (0.2) | 21 (0.2) | 4 (0.1) |  |  |
| Meeting MDD Criteria |  |  |  | <0.001 | <0.001 |
| No | 1,804 (12.1) | 1,791 (15.2) | 13 (0.4) |  |  |
| Yes | 13,093 (87.9) | 10,008 (84.8) | 3,085 (99.6) |  |  |
| Depressive Episodes |  |  |  | <0.001 | <0.001 |
| 1-2 Episodes | 1,602 (10.8) | 1,345 (11.4) | 257 (8.3) |  |  |
| 3-4 Episodes | 5,951 (39.9) | 4,384 (37.2) | 1,567 (50.6) |  |  |
| 5-6 Episodes | 3,013 (20.2) | 2,426 (20.6) | 587 (18.9) |  |  |
| 7-9 Episodes | 2,155 (14.5) | 1,664 (14.1) | 491 (15.8) |  |  |
| 10+ Episodes | 762 (5.1) | 577 (4.9) | 185 (6.0) |  |  |
| Information not provided | 1414 (9.5) | 1403 (11.9) | 11 (0.4) |  |  |
| Age of onset | 22.12±11.30 | 22.26±11.46 | 21.66±10.74 | 0.002 | 0.003 |
| Mania (ASRM) | 2.49±2.00 | 2.43±1.99 | 2.73±2.00 | <0.001 | <0.001 |
| Psychosis (CAPE) | 0.90±1.36 | 0.87±1.33 | 1.03±1.45 | <0.001 | <0.001 |
| Suicidality (SIDAS) | 8.34±9.81 | 8.06±9.70 | 9.38±10.16 | <0.001 | <0.001 |
| GAD |  |  |  | <0.001 | <0.001 |
| No | 7,543 (50.6) | 6,082 (51.5) | 1,461 (47.2) |  |  |
| Yes | 7,321 (49.1) | 5,687 (48.2) | 1,634 (52.7) |  |  |
| Information not provided | 33 (0.2) | 30 (0.3) | 3 (0.1) |  |  |
| K10 | 22.97±9.06 | 22.50±9.03 | 24.79±8.97 | <0.001 | <0.001 |
| Information not provided | 2,096 (14.1) | 1,630 (13.8) | 466 (15.0) |  |  |
| Family history* | 10,381 (69.7) | 8,180 (69.3) | 2,201 (71.0) | 0.067 | 0.079 |
| Alcohol Use Disorder |  |  |  | 0.062 | 0.077 |
| Mild | 2,328 (15.6) | 1,869 (15.8) | 459 (14.8) |  |  |
| Moderate | 1,724 (11.6) | 1,355 (11.5) | 369 (11.9) |  |  |
| Severe | 3,601 (24.2) | 2,802 (23.7) | 799 (25.8) |  |  |
| Nicotine Use Disorder |  |  |  | <0.001 | <0.001 |
| Mild | 1,453 (9.8) | 1,165 (9.9) | 288 (9.3) |  |  |
| Moderate | 1,482 (9.9) | 1,190 (10.1) | 292 (9.4) |  |  |
| Severe | 1,741 (11.7) | 1,300 (11.0) | 441 (14.2) |  |  |
| Cannabis Use Disorder |  |  |  | 0.346 | 0.390 |
| Mild | 1,033 (6.9) | 809 (6.9) | 224 (7.2) |  |  |
| Moderate | 538 (3.6) | 420 (3.6) | 118 (3.8) |  |  |
| Severe | 1,128 (7.6) | 875 (7.4) | 253 (8.2) |  |  |
| Drug Use Disorder |  |  |  | 0.001 | 0.002 |
| Mild | 1,149 (7.7) | 897 (7.6) | 252 (8.1) |  |  |
| Moderate | 642 (4.3) | 525 (4.4) | 117 (3.8) |  |  |
| Severe | 1,268 (8.5) | 955 (8.1) | 313 (10.1) |  |  |
| Medical Conditions |  |  |  |  |  |
| Diabetes or high blood sugar | 842 (5.7) | 594 (5.0) | 248 (8.0) | <0.001 | <0.001 |
| Heart attack | 136 (0.9) | 105 (0.9) | 31 (1.0) | 0.638 | 0.663 |
| Heart disease | 275 (1.8) | 220 (1.9) | 55 (1.8) | 0.800 | 0.800 |
| High blood pressure | 2,261 (15.2) | 1,697 (14.4) | 564 (18.2) | <0.001 | <0.001 |
| Stroke | 107 (0.7) | 82 (0.7) | 25 (0.8) | 0.591 | 0.635 |
| Chronic fatigue syndrome | 888 (6.0) | 681 (5.8) | 207 (6.7) | 0.063 | 0.074 |
| Chronotype (rMEQ) | 14.60±4.16 | 14.80±4.16 | 13.83±4.08 | <0.001 | <0.001 |
| Morning | 2,731 (26.4) | 2,314 (28.1) | 417 (19.9) | <0.001 | <0.001 |
| Neither | 5,067 (49.0) | 4,025 (48.8) | 1,042 (49.7) |  |  |
| Evening | 2,540 (24.6) | 1,901 (23.1) | 639 (30.5) |  |  |
| Information not provided | 4,564 (30.6) | 3,563 (30.2) | 1,001 (32.3) |  |  |
| Global Seasonality Score | 6.59±4.69 | 6.39±4.57 | 7.37±5.06 | <0.001 | <0.001 |
| ≥ Moderate seasonality | 2,190 (14.7) | 1,613 (13.7) | 577 (18.6) | <0.001 | <0.001 |
| Information not provided | 3,692 (24.8) | 2,912 (24.7) | 780 (25.2) |  |  |
| Daylight exposure workdays (h) | 1.79±1.85 | 1.84±1.89 | 1.61±1.69 | <0.001 | <0.001 |
| Information not provided | 4,471 (30.0) | 3,520 (29.8) | 951 (30.7) |  |  |
| Daylight exposure free days (h) | 3.04±2.15 | 3.16±2.18 | 2.57±1.97 | <0.001 | <0.001 |
| Information not provided | 3,761 (25.2) | 2,972 (25.2) | 789 (25.5) |  |  |

**Note:** Data are presented as Mean±Standard Deviation or n (%). Analysis of Covariance (ANCOVA) adjusting for Age and Sex was used for continuous variables; Pearson's Chi-squared was test for categorical variables. P-values are unadjusted and q-values represent false discovery rate correction for multiple testing. \*Family history indicates those with their first-degree relative(s) with any mental health disorder.
**Abbreviations:** MDD, Major Depressive Disorders; ASRM, Altman Self-Rating Mania Scale; CAPE, Community Assessment of Psychic Experience; SIDAS, Suicidal Ideation Attributes Scale; GAD, Generalized Anxiety Disorder; rMEQ, reduced Morningness-Eveningness Questionnaire.

### Polygenic risk and atypical depression

Atypical depression was significantly associated with higher MDD-PGS (odds ratio [OR], 1.10; 95%CI, 1.06-1.15; p=1.37×10^−5^), ADHD-PGS (OR, 1.08; 95%CI, 1.04-1.13; p=0.0005), BD-PGS (OR, 1.07; 95%CI, 1.02-1.12; p=0.0031), and Neuroticism-PGS (OR, 1.07; 95%CI, 1.02-1.12; p=0.0045), Figure 1A, Table S1). Atypical depression was also associated with higher PGS for BMI (OR, 1.35; 95%CI, 1.29-1.42; p=2.49×10^−37^), T2D (OR, 1.22; 95%CI, 1.16-1.28; p=8.38×10^−15^), CRP (OR, 1.12; 95%CI, 1.07-1.17; p=5.51×10^−7^), IR (OR, 1.11; 95%CI, 1.06-1.16; p=3.29×10^−6^), and lower PGS for HDL-C (OR, 0.91; 95%CI, 0.87-0.95; p=3.60×10^−5^, Figure 1B, Table S5). Atypical depression was significantly associated with lower chronotype-PGS (indicating genetic predisposition toward eveningness; OR, 0.94; 95%CI, 0.90-0.98; p=0.0055, Figure 1C, Table S9).

**Figure 1.**
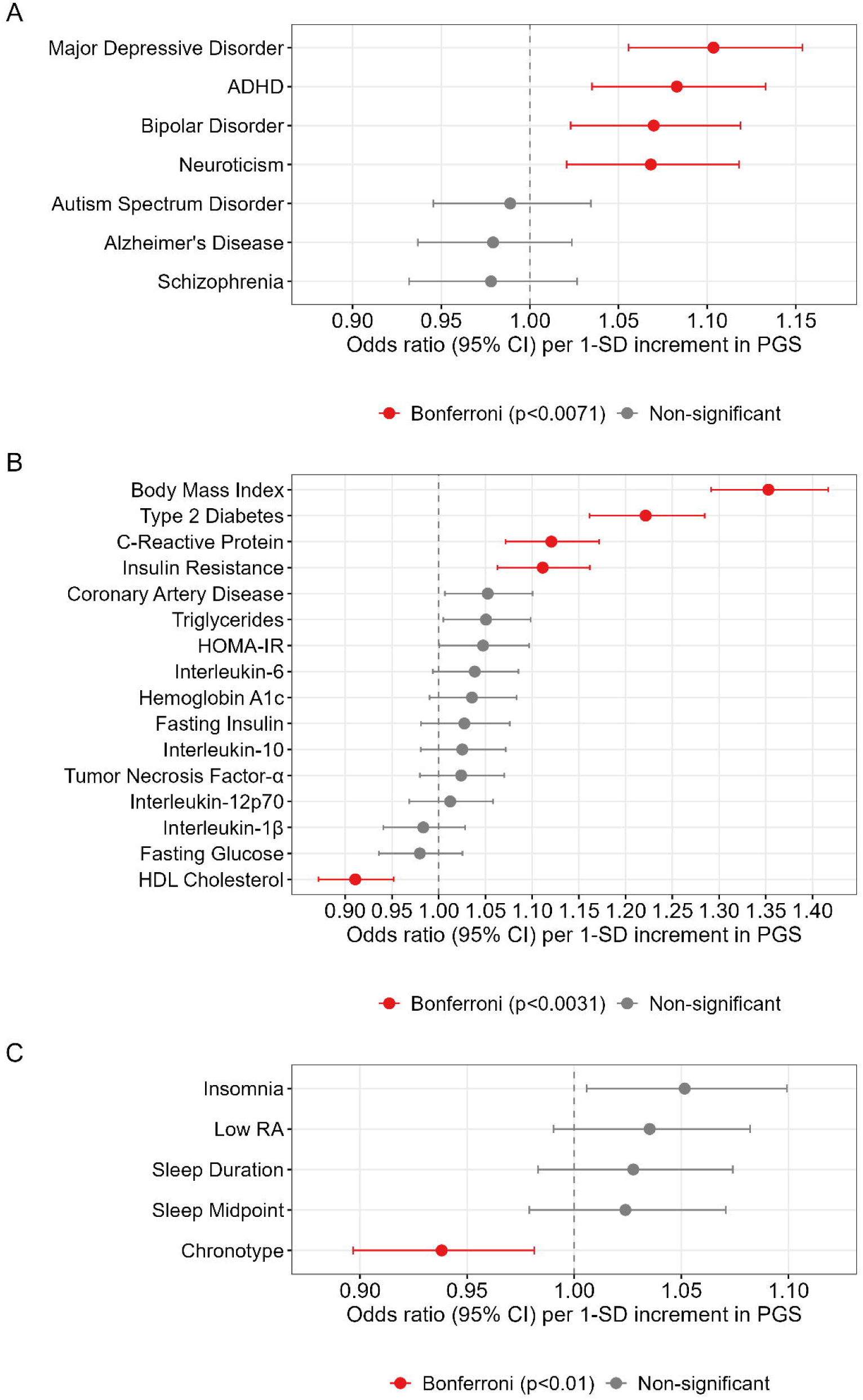
Associations between atypical depression caseness and polygenic scores (PGS) for (A) mental disorders; (B) physical health (metabolic and inflammatory-related markers); and (C) sleep and circadian-related traits (n=2,495 with atypical depression; n=9,506 with other depressive disorders). **Notes:** Results shown are each PGS with atypical depressive subtypes from separate regression models with covariates of age, sex, and the first 10 genetically-inferred ancestry PCs in logistic regression. Bars indicate 95% CI. Color-coding represents significance levels: Red: Bonferroni-corrected, and Gray: Non-significant. **Abbreviations:** ADHD, Attention-Deficit Hyperactivity Disorder; HOMA-IR, Homeostasis model assessment of insulin resistance (derived from glucose and insulin); RA, Relative amplitude.

### Treatment outcomes and atypical depression

Atypical depression was associated with poorer self-rated efficacy of SSRIs (OR, 0.88; 95% CI, 0.81-0.96; p=0.003) and SNRIs (OR, 0.85; 95% CI, 0.77-0.94; p=0.002, Figure 2, Table S13). No significant associations were found for TCA efficacy (OR, 0.93; 95% CI, 0.80-1.08). Individual antidepressant medication associations are detailed in the Supplementary Materials (Table S17).

**Figure 2.**
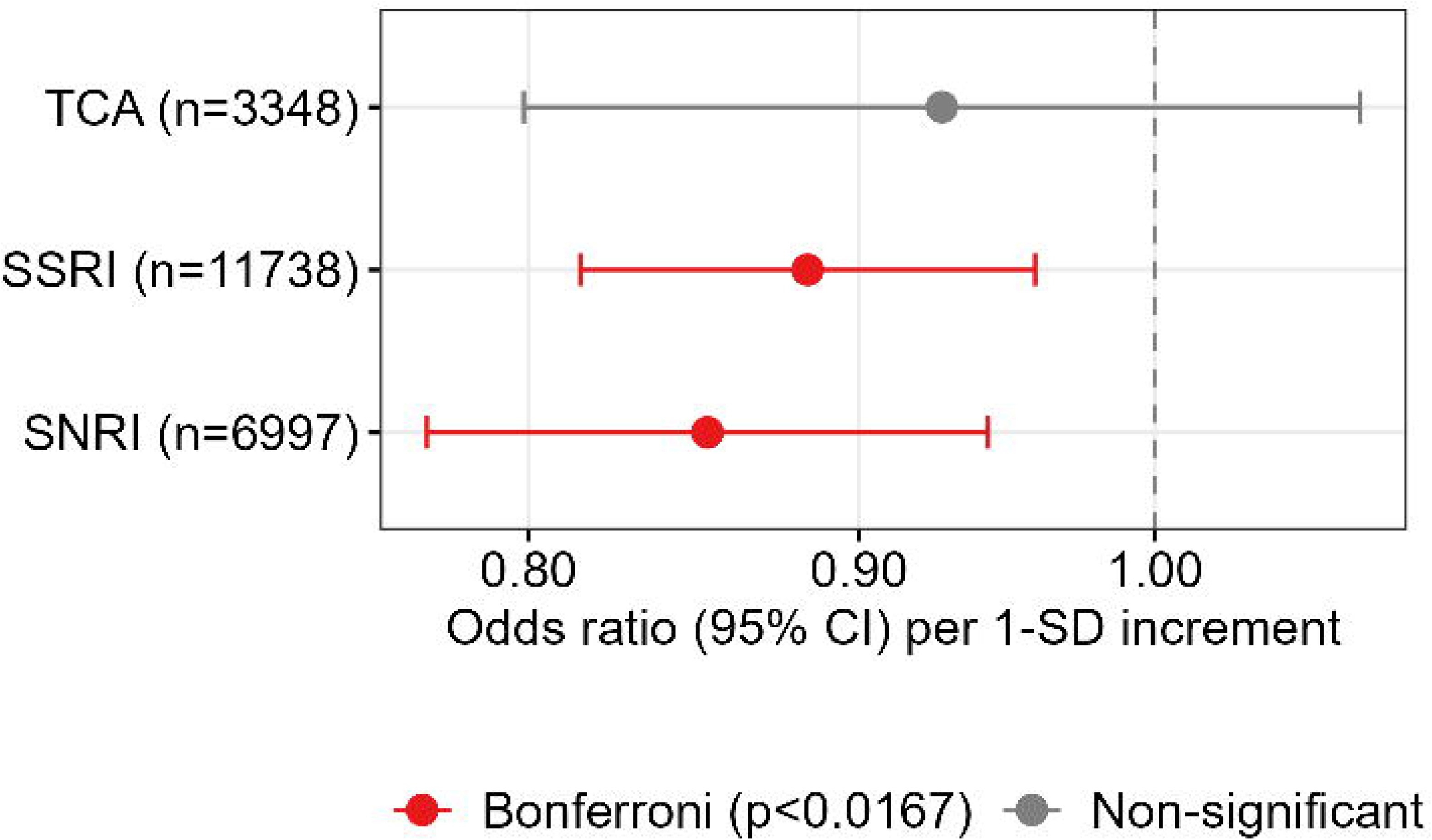
Associations of atypical depression and self-reported efficacy of antidepressants. **Note**: Results shown are each antidepressant efficacy with atypical depressive subtypes from separate regression models with covariates of age and sex in ordinal logistic regression. Bars indicate 95% CI. Color-coding represents significance levels: Red, Bonferroni-corrected, and Gray, Non-significant. **Abbreviations**: TCA, Tricyclic antidepressants; SSRI, Selective serotonin reuptake inhibitors; SNRI, Serotonin–norepinephrine reuptake inhibitor.

Twelve of the 25 queried side effects were significantly associated with atypical depression (Figure 3, Table S21). The strongest were increased likelihood of weight gain (OR, 2.89; 95%CI, 2.66-3.15; p=1.07×10^−133^) and decreased likelihood of weight loss (OR, 0.37; 95%CI, 0.28-0.48; p=5.09×10^−13^); both were expected given the atypical depression definition. Other significant side effects included drowsiness (OR=1.47), muscle pain (OR=1.41), fatigue (OR=1.39), reduced sexual desire/function (OR=1.38), dry mouth (OR=1.36), suicidal attempts (OR=1.31) and thoughts (OR=1.30), blurred vision (OR=1.28), headache (OR=1.27), and sweating (OR=1.27). No significant association was found for reporting ‘no side effects’.

**Figure 3.**
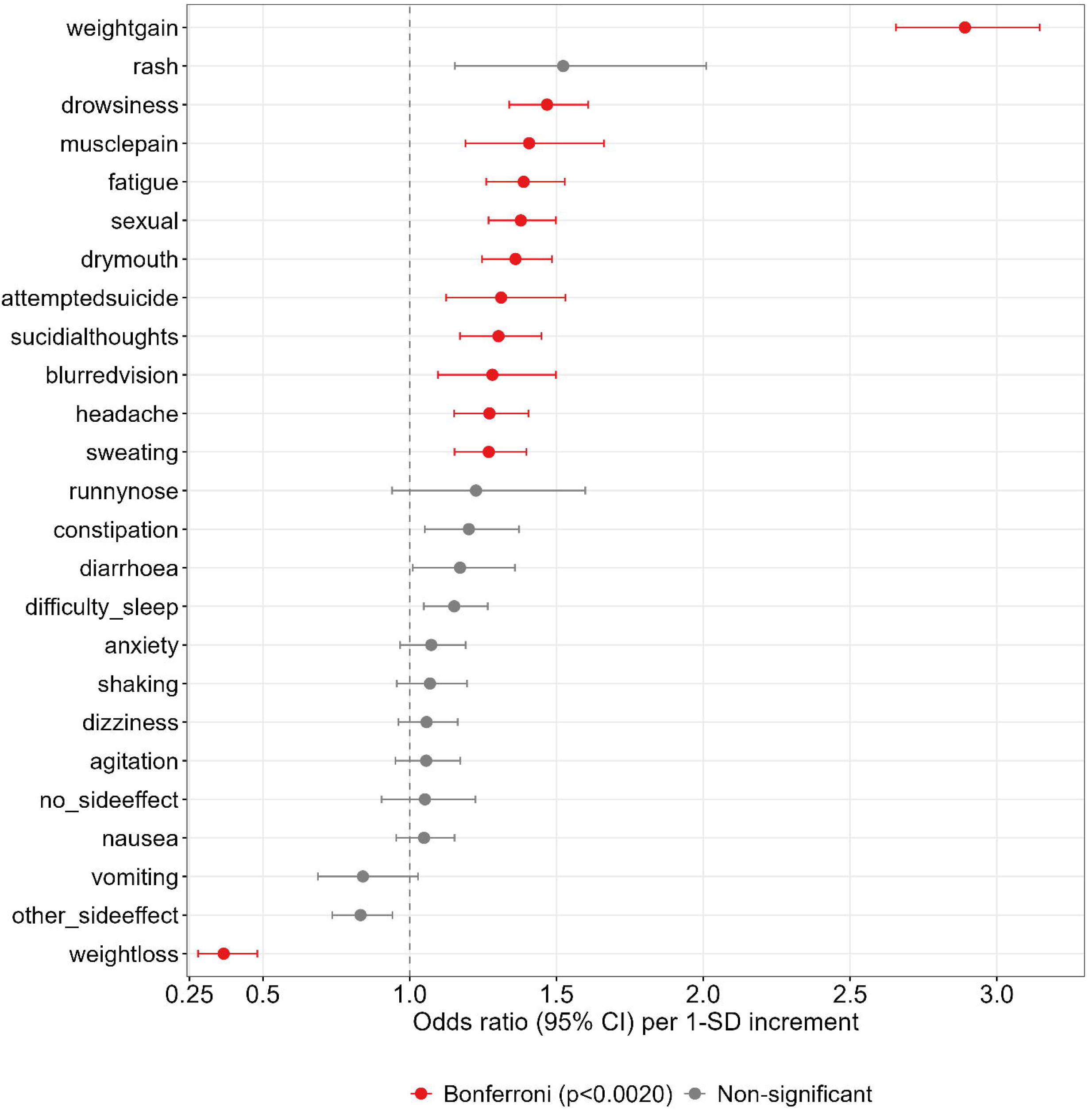
Association between atypical depression and side effects of 10 antidepressant medications (n=2,901 with atypical depression; n=10,584 with other depressive disorders). **Notes:** Results shown are each antidepressant efficacy with atypical depressive subtypes from separate regression models with covariates of age, and sex in logistic regression. Bars indicate 95% CI. Color-coding represents significance levels: Red, Bonferroni-corrected, and Gray, Non-significant.

### Sensitivity analyses

To assess the robustness of these findings, we conducted sensitivity analyses focusing on (1) participants meeting DSM-5 MDD criteria (88%); (2) sex-stratified analysis; and (3) adding BMI as an additional covariate. For MDD-confirmed cases only, similar patterns were observed with attenuated effect sizes compared to the main analysis. In sex-stratified analyses, females showed more significant associations than males (e.g., higher PGS for MDD, ADHD and BD in females vs only MDD in males), likely due to the higher female proportion in this sample (74%). The chronotype association was no longer Bonferroni significant in the sex-stratified analysis. The details are discussed in the Supplementary Materials.

When BMI was added as a covariate, no mental health or metabolic/inflammatory PGS remained significant after Bonferroni correction, though significant associations from the main analysis retained their direction (Figure 4, Table S4&S8). Notably, BMI-PGS was no longer significant (OR, 1.02, 95%CI, 0.96-1.09) when controlling for BMI itself. However, chronotype-PGS remained significant (OR, 0.92; 95%CI, 0.87-0.97; p=0.004). For treatment response, both SSRI (OR, 0.82; 95%CI, 0.74-0.91; p=0.0001) and SNRI (OR, 0.79; 95%CI, 0.70-0.89; p=0.0002) associations strengthened compared to the main analysis, whereas TCA did not survive Bonferroni correction (p=0.037). Significant side effects were reduced from 12 to eight: weight gain, drowsiness, reduced sexual function, dry mouth, fatigue, suicidal thoughts, and decreased weight loss remained significant, while constipation became newly significant (Table S24, Figure S5).

**Figure 4.**
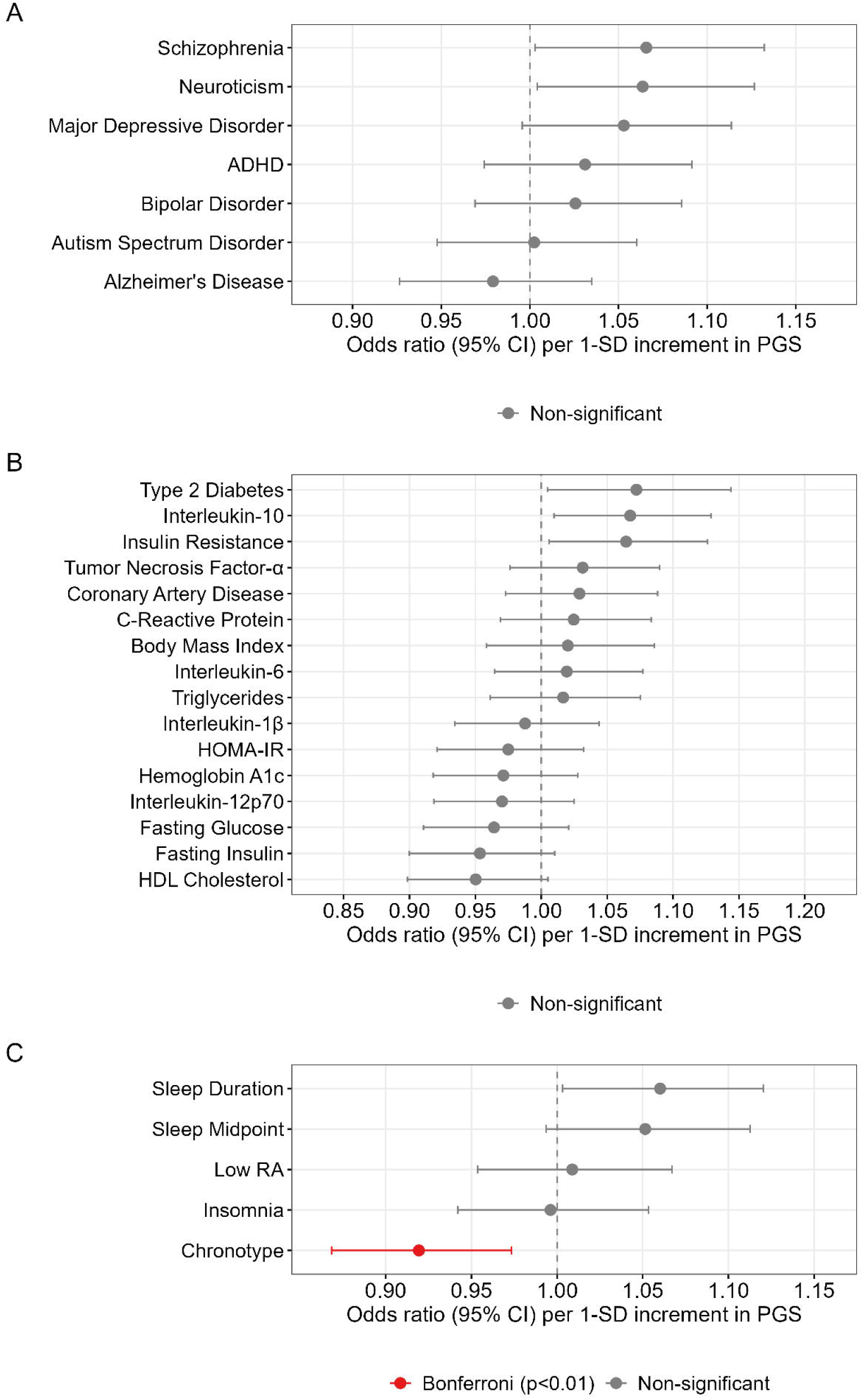
Sensitivty analysis with BMI-controlled: Associations between atypical depression caseness and polygenic scores (PGS) for (A) mental disorders; (B) physical health (metabolic and inflammatory-related markers); and (C) sleep and circadian-related traits (n=1,740 with atypical depression; n=6,511 with other depressive disorders). **Notes:** Results shown are each PGS with atypical depressive subtypes from separate regression models with covariates of age, sex, the first 10 genetically-inferred ancestry PCs, and BMI in logistic regression. Bars indicate 95% CI. Color-coding represents significance levels: Red: Bonferroni-corrected, and Gray: Non-significant. **Abbreviations:** ADHD, Attention-Deficit Hyperactivity Disorder; HOMA-IR, Homeostasis model assessment of insulin resistance (derived from glucose and insulin); RA, Relative amplitude.

## DISCUSSION

Using a large genetically-informative cohort of predominantly severe or recurrent depression,^36,42^ we identified three distinct factors supporting atypical depression’s neurobiological or clinical validity: (1) clinical characteristics including earlier onset, greater severity, higher BMI, increased comorbidity, and pronounced evening preference with less daylight exposure; (2) elevated PGS for some but not all independent mental disorders (higher PGS for MDD, ADHD, BD and Neuroticism, but not increased for autism, Alzheimer’s and schizophrenia), metabolic-inflammatory traits (higher PGS for BMI, T2D, CRP, and IR, but lower PGS for HDL-C), and circadian traits (lower PGS for chronotype); and (3) lower efficacy and more side effects from common SSRIs and SNRIs. Given the centrality of weight gain to the case definition of atypical depression, sensitivity analysis controlling for BMI revealed that mental health and metabolic-inflammatory PGS associations disappeared, whereras chronotype-PGS and treatment response differences persisted, suggesting BMI-mediated and BMI-independent pathways.

Our case definition prioritized reversed neurovegetative symptoms (hypersomnia and weight gain) due to their stronger and more consistent biological associations than other atypical features.^10,11^ In severe, recurrent depression (characteristic of this sample) neurovegetative symptoms tend to be more stable across episodes.^12,13^ Consistent with previous research, atypical depression cases were more likely to be female, with earlier onset, more depressive episodes, higher suicidality, and greater comorbidity with anxiety, psychotic, and manic features.^12,15,25,53,54^ We also found more severe nicotine and drug use disorders in atypical depression, extending prior findings of high smoking rates across mental disorders to show subtype-specific substance use vulnerabilities.^55^ Notably, we identified stronger evening preference, greater seasonality, and significantly less time in daylight, suggesting circadian disruption as a potential biological marker.

Previous AGDS reports demonstrated differential genetic associations across depressive subtypes.^43^ We build on this using a larger sample, analyzing metabolic, inflammatory and circadian PGS, assessing treatment response differences, employing updated PGS scoring methods with better-powered GWAS, and examining sex-stratified and BMI-adjusted associations. Four mental health PGS (ADHD, MDD, BD, Neuroticism) showed significant associations with atypical depression. These findings align with clinical evidence showing higher atypical feature rates in BD type-II or bipolar spectrum disorder.^56,57^ The MDD-PGS association may reflect atypical depression’s greater severity and functional impairment^12,13^, while ADHD- and Neurotism-PGS imply neurodevelopmental and emotional regulation pathways, consistent with heightened emotional reactivity and impulsivity.^1,56,58–60^ A previous AGDS study reported an MDD-PGS association with dimensional somatic symptoms related to atypical features.^51^ Genetic associations with metabolic-inflammatory traits further validate the biologic basis of atypical depression.^10,11,29,38,42,61,62^ Notably, the association was observed with IR-PGS (derived from Triglyceride:HDL-C ratio) but not HOMA-IR-PGS (derived from glucose and insulin), possibly reflecting different insulin resistance aspects or weaker genetic instruments for HOMA-IR-PGS.^63^ However, genetic associations with metabolic traits should be interpreted cautiously given that our atypical depression definition includes weight gain as one of two central features.

In BMI-covariate sensitivity analyses, all mental health and metabolic-inflammatory PGS associations became non-significant, potentially reflecting either BMI’s mediating role or reduced statistical power due to substantial BMI missingness (31%). While our cross-sectional design limits causal inference, previous research provides relevant context. Mendelian randomization evidence suggests that higher BMI causally influences increased appetite (an atypical feature) but not atypical depression as a whole.^62^ Conversely, prospective data show that atypical depression predicts subsequent weight gain and metabolic dysfunction.^64^ These suggest complex bidirectional relationships between BMI and atypical depression.

Notably, chronotype-PGS remained robust after BMI adjustment, suggesting that circadian-related genetic factors contribute to atypical depression through pathways not fully captured by BMI alone. This finding, combined with observed evening preference and reduced daylight exposure, provides evidence for circadian disruption as a core feature of atypical depression.^40,65^

Consistent with our hypothesis, atypical depression was associated with lower SSRI and SNRI efficacy, and more side effects. Critically, treatment differences strengthened after BMI adjustment, demonstrating that poor response extends beyond weight-related factors. This aligns with another AGDS study showing that phenotypic eveningness was associated with lower self-reported antidepressant efficacy and more side effects.^66^ The side effect profile in atypical depression remained largely unchanged after BMI adjustment. Research suggests that antidepressant side effects may share common underlying factors (e.g., genetic liability to higher BMI or insomnia) rather than being drug-specific.^67^ The persistence of both chronotype-PGS associations and treatment resistance after BMI adjustment suggests circadian disruption may be a key mechanism underlying treatment resistance in atypical depression.^40^ This is further supported by research showing that later sleep midpoint and reduced physical activity partially mediated the relationship between atypical depression and elevated BMI and metabolic syndrome.^68^ These findings suggest that people with atypical depression may benefit from: (1) earlier consideration of alternatives to SSRIs/SNRI; and (2) circadian-targeted interventions as adjunctive treatments. The BMI-independent chronotype associations and treatment resistance patterns indicate that “circadian medicine” approaches may be particularly relevant for this population, potentially improving both sleep-wake regulation and antidepressant response.^69–71^

Our study has several limitations. First, while our atypical depression definition using weight gain and hypersomnia aligns with established criteria focused on reversed neurovegetative phenomena, this may introduce circularity when interpreting genetic associations with BMI. Second, the substantial BMI missingness (31%) limits interpretation of covariate analyses. The cross-sectional design prevents distinguishing between mediation, confounding, and reverse causation in genetic associations. Third, the antidepressants explored and the binary classification of depression subtypes may oversimplify heterogeneity. Finally, our sample is limited to individuals with genetically-inferred European ancestry, which may reduce generalizability.

Altogether, this study provides evidence that atypical depression represents a clinically meaningful subtype predicting differential treatment response patterns and polygenic risk profiles, some of which are independent of BMI status. The robust chronotype-PGS association and persistent treatment resistance after BMI adjustment support the concept that circadian disruption is a key pathway warranting targeted interventions. While genetic associations with mental disorder and metabolic traits largely reflected BMI-related variance, the BMI-independent findings support investigating circadian-based treatments and alternative pharmacological approaches for this depressive subtype.

## Data Availability

All data produced in the present study are available upon reasonable request to the authors

## Acknowledgements

We are indebted to all of the Australian Genetics of Depression Study participants for giving their time to contribute to this study. We thank all the people who helped in the conception, implementation, beta testing, media campaign, and data cleaning. The study protocol used by 23andMe, Inc., was approved by an external AAHRPP-accredited institutional review board. We would like to thank the research participants and employees of 23andMe, Inc., for making this work possible. The GWAS summary statistics from Howard et al. include those provided by 23andMe, Inc. The Australian Genetics of Depression Study was primarily funded by grant 1086683 from the National Medical Health and Research Council (NHMRC) of Australia. This work was further supported by philanthropic donations from families who are affected by mental illness (who would like to be left anonymous) awarded to MS; NHMRC EL1 Investigator Grants awarded to JJC and BLM (GNT2008196 and GNT2017176, respectively); a Breakthrough Mental Health Research Foundation Fellowship awarded to ET; an NHMRC L1 Investigator Grant (APP117291) awarded to SEM; and an NHMRC L3 Investigator Grant (GNT2016346) awarded to IBH.

## References

1. West ED, Dally PJ. Effects of iproniazid in depressive syndromes. Br Med J. 1959;1(5136):1491–1494.

2. American Psychiatric Association. Diagnostic and statistical manual of mental disorders: DSM-5. Vol 5: American psychiatric association Washington, DC; 2013.

3. Thase ME. Recognition and diagnosis of atypical depression. J Clin Psychiatry. 2007;68 Suppl 8:11–16.

4. Thase ME. Atypical Depression: Useful Concept, but it’s Time to Revise the DSM-IV Criteria. Neuropsychopharmacology. 2009;34(13):2633–2641.

5. Stewart JW, McGrath PJ, Rabkin JG, Quitkin FM. Atypical Depression: A Valid Clinical Entity? Psychiatric Clinics of North America. 1993;16(3):479–495.

6. Parker G, Roy K, Mitchell P, Wilhelm K, Malhi G, Hadzi-Pavlovic D. Atypical depression: a reappraisal. Am J Psychiatry. 2002;159(9):1470–1479.

7. Posternak MA, Zimmerman M. Partial Validation of the Atypical Features Subtype of Major Depressive Disorder. Archives of General Psychiatry. 2002;59(1):70–76.

8. Parker G, Parker K, Mitchell P, Wilhelm K. Atypical depression: Australian and US studies in accord - special commentary. Current Opinion in Psychiatry. 2005;18(1):1–5.

9. Quitkin F, McGrath P, Stewart J, et al. Phenelzine and imipramine in mood reactive depressives. Further delineation of the syndrome of atypical depression. Journal of Clinical Psychopharmacology. 1990;10(2):138.

10. Milaneschi Y, Lamers F, Peyrot WJ, et al. Polygenic dissection of major depression clinical heterogeneity. Molecular Psychiatry. 2016;21(4):516–522.

11. Milaneschi Y, Lamers F, Peyrot WJ, et al. Genetic Association of Major Depression With Atypical Features and Obesity-Related Immunometabolic Dysregulations. JAMA Psychiatry. 2017;74(12):1214–1225.

12. Brailean A, Curtis J, Davis K, Dregan A, Hotopf M. Characteristics, comorbidities, and correlates of atypical depression: evidence from the UK Biobank Mental Health Survey. Psychol Med. 2020;50(7):1129–1138.

13. Vreijling SR, van Haeringen M, Milaneschi Y, et al. Sociodemographic, lifestyle and clinical characteristics of energy-related depression symptoms: A pooled analysis of 13,965 depressed cases in 8 Dutch cohorts. Journal of Affective Disorders. 2023;323:1–9.

14. Benazzi F. Can only reversed vegetative symptoms define atypical depression? European Archives of Psychiatry and Clinical Neuroscience. 2002;252:288–293.

15. Matza LS, Revicki DA, Davidson JR, Stewart JW. Depression With Atypical Features in the National Comorbidity Survey: Classification, Description, and Consequences. Archives of General Psychiatry. 2003;60(8):817–826.

16. Lee S, Ng KL, Tsang A. Prevalence and Correlates of Depression with Atypical Symptoms in Hong Kong. Australian & New Zealand Journal of Psychiatry. 2009;43(12):1147–1154.

17. Pande AC, Birkett M, Fechner-Bates S, Haskett RF, Greden JF. Fluoxetine versus phenelzine in atypical depression. Biological Psychiatry. 1996;40(10):1017–1020.

18. SÖgaard J, Lane R, Latimer P, et al. A 12-week study comparing moclobemide and sertraline in the treatment of outpatients with atypical depression. Journal of Psychopharmacology. 1999;13(4):406–414.

19. Lonnqvist J, Sihvo S, Syvälahti E, Kiviruusu O. Moclobemide and fluoxetine in atypical depression: a double-blind trial. Journal of affective disorders. 1994;32(3):169–177.

20. McGrath PJ, Stewart JW, Janal MN, Petkova E, Quitkin FM, Klein DF. A Placebo-Controlled Study of Fluoxetine Versus Imipramine in the Acute Treatment of Atypical Depression. American Journal of Psychiatry. 2000;157(3):344–350.

21. Henkel V, Mergl R, Allgaier A-K, Kohnen R, Möller H-J, Hegerl U. Treatment of depression with atypical features: A meta-analytic approach. Psychiatry Research. 2006;141(1):89–101.

22. Suchting R, Tirumalaraju V, Gareeb R, et al. Revisiting monoamine oxidase inhibitors for the treatment of depressive disorders: A systematic review and network meta-analysis. Journal of Affective Disorders. 2021;282:1153–1160.

23. Nierenberg AA, Alpert JE, Pava J, Rosenbaum JF, Fava M. Course and treatment of atypical depression. Journal of Clinical Psychiatry. 1998;59(18):5–9.

24. Agosti V, Stewart JW. Atypical and non-atypical subtypes of depression: comparison of social functioning, symptoms, course of illness, co-morbidity and demographic features. Journal of affective disorders. 2001;65(1):75–79.

25. Blanco C, Vesga-López O, Stewart JW, Liu S-M, Grant BF, Hasin DS. Prevalence, correlates, comorbidity and treatment-seeking among individuals with a lifetime major depressive episode with and without atypical features: results from the National Epidemiologic Survey on alcohol and related conditions. The Journal of clinical psychiatry. 2012;73(2):224.

26. Withers AC, Tarasoff JM, Stewart JW. Is depression with atypical features associated with trauma history? The Journal of clinical psychiatry. 2013;74(5):9897.

27. Fries E, Hesse J, Hellhammer J, Hellhammer DH. A new view on hypocortisolism. Psychoneuroendocrinology. 2005;30(10):1010–1016.

28. Newport DJ, Heim C, Bonsall R, Miller AH, Nemeroff CB. Pituitary-adrenal responses to standard and low-dose dexamethasone suppression tests in adult survivors of child abuse. Biol Psychiatry. 2004;55(1):10–20.

29. Lamers F, Vogelzangs N, Merikangas K, De Jonge P, Beekman A, Penninx B. Evidence for a differential role of HPA-axis function, inflammation and metabolic syndrome in melancholic versus atypical depression. Molecular psychiatry. 2013;18(6):692–699.

30. Lamers F, Milaneschi Y, Vinkers CH, Schoevers RA, Giltay EJ, Penninx BWJH. Depression profilers and immuno-metabolic dysregulation: Longitudinal results from the NESDA study. Brain, Behavior, and Immunity. 2020;88:174–183.

31. Pan A, Keum N, Okereke OI, et al. Bidirectional association between depression and metabolic syndrome: a systematic review and meta-analysis of epidemiological studies. Diabetes Care. 2012;35(5):1171–1180.

32. Vancampfort D, Stubbs B, Mitchell AJ, et al. Risk of metabolic syndrome and its components in people with schizophrenia and related psychotic disorders, bipolar disorder and major depressive disorder: a systematic review and meta-analysis. World Psychiatry. 2015;14(3):339–347.

33. Silva DA, Coutinho E, Ferriani LO, Viana MC. Depression subtypes and obesity in adults: A systematic review and meta-analysis. Obes Rev. 2020;21(3):e12966.

34. Shell AL, Crawford CA, Cyders MA, Hirsh AT, Stewart JC. Depressive disorder subtypes, depressive symptom clusters, and risk of obesity and diabetes: A systematic review. J Affect Disord. 2024;353:70–89.

35. Bunker SJ, Colquhoun DM, Esler MD, et al. “Stress” and coronary heart disease: psychosocial risk factors. Medical Journal of Australia. 2003;178(6):272–276.

36. Nguyen TD, Harder A, Xiong Y, et al. Genetic heterogeneity and subtypes of major depression. Mol Psychiatry. 2022;27(3):1667–1675.

37. Scott EM, Carpenter JS, Iorfino F, et al. What is the prevalence, and what are the clinical correlates, of insulin resistance in young people presenting for mental health care? A cross-sectional study. BMJ open. 2019;9(5):e025674.

38. Penninx BWJH, Milaneschi Y, Lamers F, Vogelzangs N. Understanding the somatic consequences of depression: biological mechanisms and the role of depression symptom profile. BMC Medicine. 2013;11(1):129.

39. Tickell AM, Rohleder C, Ho N, et al. Identifying pathways to early-onset metabolic dysfunction, insulin resistance and inflammation in young adult inpatients with emerging affective and major mood disorders. Early Intervention in Psychiatry. 2022;16(10):1121–1129.

40. Carpenter JS, Crouse JJ, Scott EM, et al. Circadian depression: A mood disorder phenotype. Neurosci Biobehav Rev. 2021;126:79–101.

41. Hickie IB, Naismith SL, Robillard R, Scott EM, Hermens DF. Manipulating the sleep-wake cycle and circadian rhythms to improve clinical management of major depression. BMC Med. 2013;11:79.

42. Badini I, Coleman JRI, Hagenaars SP, et al. Depression with atypical neurovegetative symptoms shares genetic predisposition with immuno-metabolic traits and alcohol consumption. Psychol Med. 2022;52(4):726–736.

43. Mitchell BL, Campos AI, Whiteman DC, et al. The Australian Genetics of Depression Study: New Risk Loci and Dissecting Heterogeneity Between Subtypes. Biol Psychiatry. 2022;92(3):227–235.

44. Oliva V, Fanelli G, Kasper S, et al. Melancholic features and typical neurovegetative symptoms of major depressive disorder show specific polygenic patterns. Journal of Affective Disorders. 2023;320:534–543.

45. Byrne EM, Kirk KM, Medland SE, et al. Cohort profile: the Australian genetics of depression study. BMJ open. 2020;10(5):e032580.

46. Purcell S, Neale B, Todd-Brown K, et al. PLINK: a tool set for whole-genome association and population-based linkage analyses. Am J Hum Genet. 2007;81(3):559–575.

47. Chang CC, Chow CC, Tellier LC, Vattikuti S, Purcell SM, Lee JJ. Second-generation PLINK: rising to the challenge of larger and richer datasets. Gigascience. 2015;4:7.

48. Taliun D, Harris DN, Kessler MD, et al. Sequencing of 53,831 diverse genomes from the NHLBI TOPMed Program. Nature. 2021;590(7845):290–299.

49. Jiang L, Zheng Z, Qi T, et al. A resource-efficient tool for mixed model association analysis of large-scale data. Nature Genetics. 2019;51(12):1749–1755.

50. Zheng Z, Liu S, Sidorenko J, et al. Leveraging functional genomic annotations and genome coverage to improve polygenic prediction of complex traits within and between ancestries. Nature Genetics. 2024;56(5):767–777.

51. Mitchell BL, Thorp JG, Wu Y, et al. Polygenic Risk Scores Derived From Varying Definitions of Depression and Risk of Depression. JAMA Psychiatry. 2021;78(10):1152–1160.

52. R: A Language and Environment for Statistical Computing [computer program]. Vienna, Austria: R Foundation for Statistical Computing; 2022.

53. Novick JS, Stewart JW, Wisniewski SR, et al. Clinical and Demographic Features of Atypical Depression in Outpatients With Major Depressive Disorder: Preliminary Findings From STAR? D. Journal of Clinical Psychiatry. 2005;66(8):1002–1011.

54. Xin L-M, Chen L, Su Y-A, et al. Prevalence and clinical features of atypical depression among patients with major depressive disorder in China. Journal of Affective Disorders. 2019;246:285–289.

55. Glaus J, Vandeleur C, Gholam-Rezaee M, et al. Atypical depression and alcohol misuse are related to the cardiovascular risk in the general population. Acta Psychiatr Scand. 2013;128(4):282–293.

56. Perugi G, Fornaro M, Akiskal HS. Are atypical depression, borderline personality disorder and bipolar II disorder overlapping manifestations of a common cyclothymic diathesis? World Psychiatry. 2011;10(1):45–51.

57. Łojko D, Rybakowski JK. Atypical depression: current perspectives. Neuropsychiatr Dis Treat. 2017;13:2447–2456.

58. Davidson JR, Miller RD, Turnbull CD, Sullivan JL. Atypical depression. Arch Gen Psychiatry. 1982;39(5):527–534.

59. Angst J, Gamma A, Sellaro R, Zhang H, Merikangas K. Toward validation of atypical depression in the community: results of the Zurich cohort study. J Affect Disord. 2002;72(2):125–138.

60. Chopra KK, Bagby RM, Dickens S, Kennedy SH, Ravindran A, Levitan RD. A dimensional approach to personality in atypical depression. Psychiatry Res. 2005;134(2):161–167.

61. Penninx BWJH. Depression and cardiovascular disease: Epidemiological evidence on their linking mechanisms. Neuroscience & Biobehavioral Reviews. 2017;74:277–286.

62. Pistis G, Milaneschi Y, Vandeleur CL, et al. Obesity and atypical depression symptoms: findings from Mendelian randomization in two European cohorts. Translational Psychiatry. 2021;11(1):96.

63. Dupuis J, Langenberg C, Prokopenko I, et al. New genetic loci implicated in fasting glucose homeostasis and their impact on type 2 diabetes risk. Nat Genet. 2010;42(2):105–116.

64. Lasserre AM, Glaus J, Vandeleur CL, et al. Depression with atypical features and increase in obesity, body mass index, waist circumference, and fat mass: a prospective, population-based study. JAMA Psychiatry. 2014;71(8):880–888.

65. Crouse JJ, Carpenter JS, Song YJC, et al. Circadian rhythm sleep-wake disturbances and depression in young people: implications for prevention and early intervention. Lancet Psychiatry. 2021;8(9):813–823.

66. Crouse JJ, Park SH, Byrne EM, et al. Evening Chronotypes With Depression Report Poorer Outcomes of Selective Serotonin Reuptake Inhibitors: A Survey-Based Study of Self-Ratings. Biological Psychiatry. 2024;96(1):4–14.

67. Campos AI, Mulcahy A, Thorp JG, et al. Understanding genetic risk factors for common side effects of antidepressant medications. Communications Medicine. 2021;1(1):45.

68. Glaus J, Kang SJ, Guo W, et al. Objectively assessed sleep and physical activity in depression subtypes and its mediating role in their association with cardiovascular risk factors. J Psychiatr Res. 2023;163:325–336.

69. Hickie IB, Crouse JJ. Sleep and circadian rhythm disturbances: plausible pathways to major mental disorders? World Psychiatry. 2024;23(1):150–151.

70. Panda S. The arrival of circadian medicine. Nat Rev Endocrinol. 2019;15(2):67–69.

71. Hickie IB, Rogers NL. Novel melatonin-based therapies: potential advances in the treatment of major depression. Lancet. 2011;378(9791):621–631.

