## supplementary materials for "Atypical depression is associated with a distinct clinical, neurobiological, treatment response and polygenic risk profile"

**Questionnaires we examined in this study:**

- *DSM-5 criteria for Major Depressive Disorder* (MDD)(1): Diagnostic criteria for MDD.
- *DSM-5 criteria for Generalized Anxiety Disorder* (GAD)(1): Diagnostic criteria for GAD.
- *DSM-5 criteria for Substance Use Disorder* (1): Diagnostic criteria for Alcohol Use Disorder, Nicotine Use Disorder (combining tobacco and e-cigarette use), Cannabis Use Disorder, and Drug Use Disorder (including Cocaine, Amphetamine-type stimulants, Inhalants, Sedatives/sleeping pills, Hallucinogens, Opioids, Ecstasy, Ketamine, GHB, Other party drugs)
- *Altman Self-Rating Mania Scale* (ASRM)(2): 5-item scale assessing manic symptoms (higher scores indicate a higher probability of a manic or hypomanic condition).
- *Community Assessment of Psychic Experiences* (CAPE) (modified version)(3): 6-item scale assessing psychotic-like symptoms (higher scores indicate higher severity of psychotic-like experiences).
- *Suicidal Ideation Attributes Scale* (SIDAS)(4): 5-item questionnaire evaluating suicidal ideation attributes (higher scores indicate more severe suicidal thoughts).
- *Kessler Psychological Distress scale* (K10)(5): 10-item scale measuring psychological distress (higher scores indicate higher distress).
- *Morningness-Eveningness Questionnaire*, reduced version (rMEQ)(6): 5-item questionnaire (higher scores indicate greater tendency toward morningness).
- *Outdoor light exposure:* Participants reported their average daily time spent in natural light, separately for workdays and free days (e.g., weekend), measured in hours and minutes.
- *Seasonal Pattern Assessment Questionnaire* (SPAQ) (7)*:* 17-item self-report questionnaire developed to assess seasonality in mood disorder patients as well as the general population. We considered six items measuring the degree of seasonal change in sleep length, social activity, mood, weight, appetite, and energy, on a scale from 0 (no change) to 4 (extremely marked change). Item scores were summed to derive the General Seasonality Score (GSS; scored 0 – 24).
- *Antidepressant Efficacy:* Efficacy of each antidepressant was assessed with the question: “How well does/did each antidepressant work for you?” The responses were analyzed on the ordinal scale: not at all well (0), moderately well (1), very well (2) (“I don’t know” responses were excluded here). Efficacy was compared across three major antidepressant classes: SSRIs (sertraline, escitalopram, citalopram, fluoxetine, paroxetine), SNRIs (venlafaxine, desvenlafaxine, duloxetine), and TCAs (amitriptyline, mirtazapine).
- *Side effects*: Side effects were assessed with the question: “Which side effects did you experience from the following antidepressant(s)?” Participants were only asked to report side effects if they had taken a given antidepressant. The following were queried: dry mouth, sweating, nausea, vomiting, diarrhea, constipation, headache, dizziness, shaking, muscle pain, drowsiness, difficulty getting to sleep, increased anxiety, agitation, fatigue or weakness, weight gain, weight loss, rash, runny nose, reduced sexual desire/function, blurred vision, suicidal thoughts, attempted suicide, other side effects, and no side effects. Responses were analyzed as a binary variable: no (0) or yes (1).

**Table S1. Polygenic risk scores (PGS) examined.**

| **Trait** | **GWAS citation** |
| --- | --- |
| **Psychiatric/neurological** |  |
| ADHD | (8) |
| Alzheimer’s disease | (9) |
| Autism | (10) |
| Bipolar disorder | (11) |
| Major depressive disorder | (12) |
| Neuroticism | (13) |
| Schizophrenia | (14) |
| **Metabolic** |  |
| Body mass index | (15) |
| Coronary artery disease | (16) |
| Fasting insulin | (17) |
| Fasting glucose | (18) |
| Glycated hemoglobin (HbA1c) | (19) |
| HDL-Cholesterol | (19) |
| Insulin Resistance | (20) |
| Homeostatic Model Assessment  of Insulin Resistance (HOMAIR) | (17) |
| Triglycerides | (19) |
| Type 2 diabetes | (21) |
| **Inflammatory** |  |
| C-reactive protein | (22) |
| Interleukin-6 (IL-6) | (23) |
| Interleukin-10 (IL-10) | (23) |
| Interleukin-12p70 (IL-12) | (23) |
| Interleukin-1β (IL-1β) | (23) |
| Tumor necrosis factor α (TNF-α) | (23) |
| **Sleep-circadian traits** |  |
| Sleep midpoint | (24) |
| Relative amplitude | (25) |
| Sleep duration | (26) |
| Insomnia | (26) |
| Chronotype | (24) |

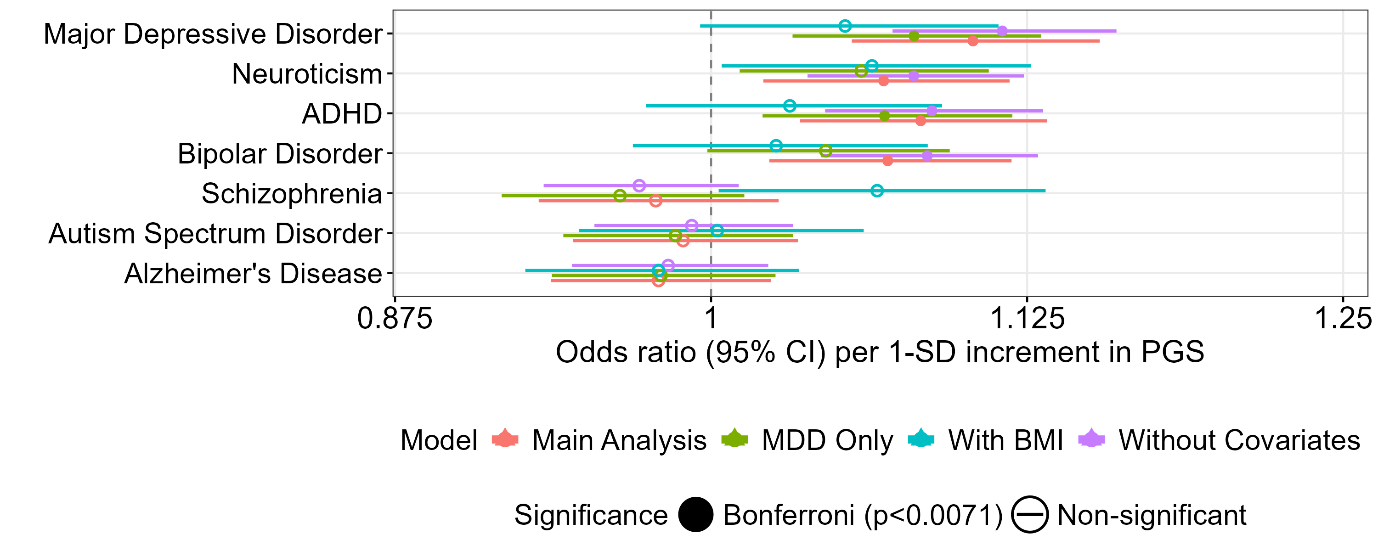

**Figure S1. Association between atypical depression caseness and polygenic scores (PGS) for mental disorders: main analysis and sensitivity analyses**

**Notes:** Sensitivity analyses included: (1) BMI-adjusted analysis (n=8,251) compared to main analysis (n=12,001) controlling for age, sex, BMI, and first 10 genetically-inferred ancestry PCs; (2) MDD-confirmed cases only (n=10,511), controlling for age, sex, and first 10 genetically-inferred ancestry PCs; (3) unadjusted model (n=14,897). Results show odds ratios from separate logistic regression models for each PGS with Bonferroni correction. Bars indicate 95% CI.

**Abbreviations:** ADHD, Attention-Deficit Hyperactivity Disorder

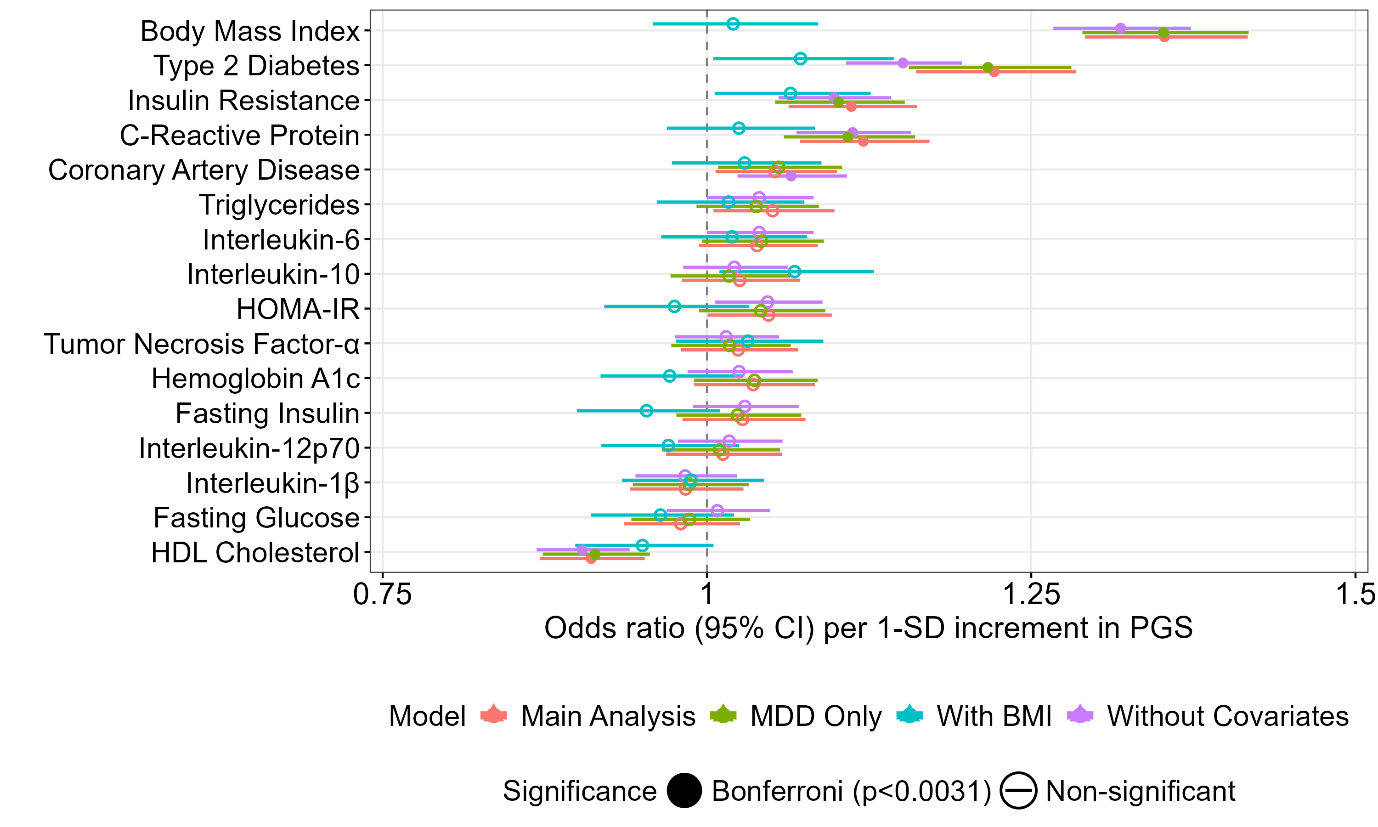
**Figure S2. Association between atypical depression and polygenic scores (PGS) for metabolic and inflammatory traits: main analysis and sensitivity analyses**

**Notes:** Sensitivity analyses included: (1) BMI-adjusted analysis (n=8,251) compared to main analysis (n=12,001) controlling for age, sex, BMI and first 10 genetically-inferred ancestry PCs; (2) MDD-confirmed cases only (n=10,511), controlling for age, sex, and first 10 genetically-inferred ancestry PCs; (3) unadjusted model (n=14,897). Results show odds ratios from separate logistic regression models for each PGS with Bonferroni correction. Bars indicate 95% CI.

**Abbreviations:** HOMA-IR, Homeostasis model assessment of insulin resistance (derived from glucose and insulin)

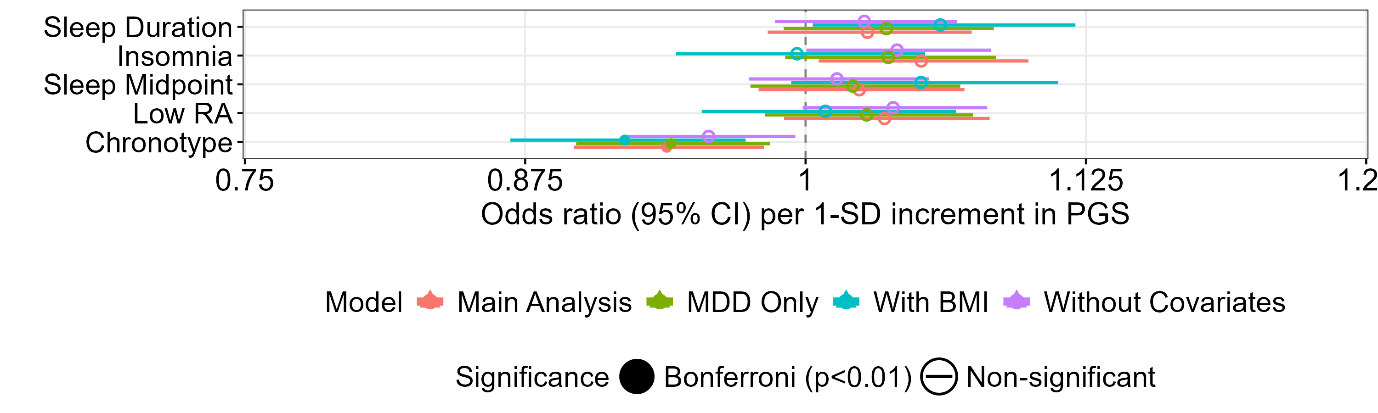

**Figure S3. Association between atypical depression and polygenic scores for sleep/circadian-related traits: main analysis and sensitivity analyses**

**Notes:** Sensitivity analyses included: (1) BMI-adjusted analysis (n=8,251) compared to main analysis (n=12,001) controlling for age, sex, BMI and first 10 genetically-inferred ancestry PCs; (2) MDD-confirmed cases only (n=10,511), controlling for age, sex, and first 10 genetically-inferred ancestry PCs; (3) unadjusted model (n=14,897). Results show odds ratios from separate logistic regression models for each PGS with Bonferroni correction. Bars indicate 95% CI.

**Abbreviations:** RA, Relative amplitude

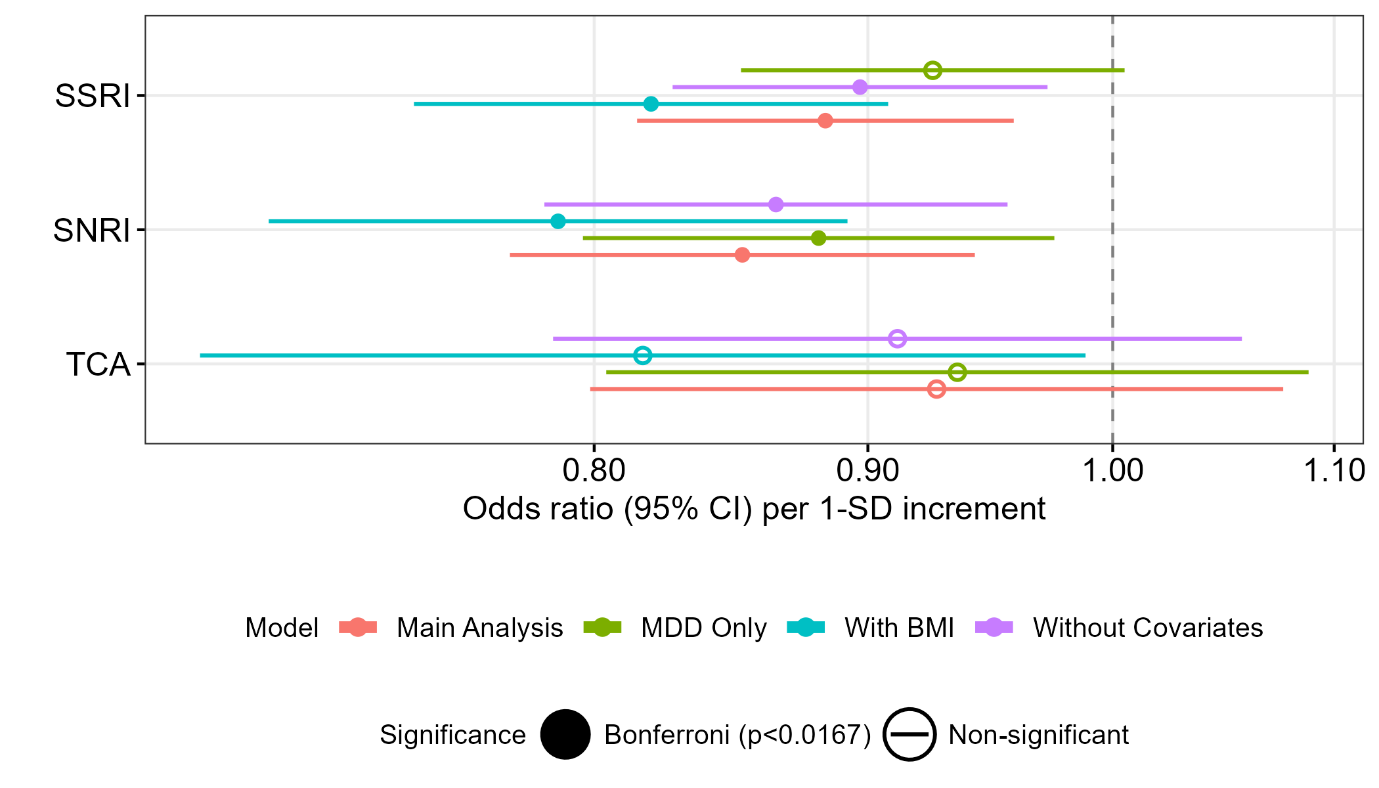

**Figure S4. Association between atypical depression and self-reported efficacy of antidepressants: main analysis and sensitivity analyses**

**Notes:** Sensitivity analyses included: (1) BMI-adjusted analysis (SSRI, n=8,122; SNRI, n=4,819; TCA, n=2,226) compared to main analysis controlling for age, sex and BMI (SSRI, n=11,738; SNRI, n=6,997; TCA, n=3,348); (2) MDD-confirmed cases only (SSRI, n=10,507; SNRI, n=6,439; TCA, n=3,084); (3) unadjusted model (SSRI, n=11,747; SNRI, n=7,000; TCA, n=3,351). Results show odds ratios from separate ordinal regression models for each PGS with Bonferroni correction. Bars indicate 95% CI.

**Abbreviations:** SSRI, Selective Serotonin Reuptake Inhibitors; SNRI, Serotonin-Norepinephrine Reuptake Inhibitors; TCA, Tricyclic Antidepressants.

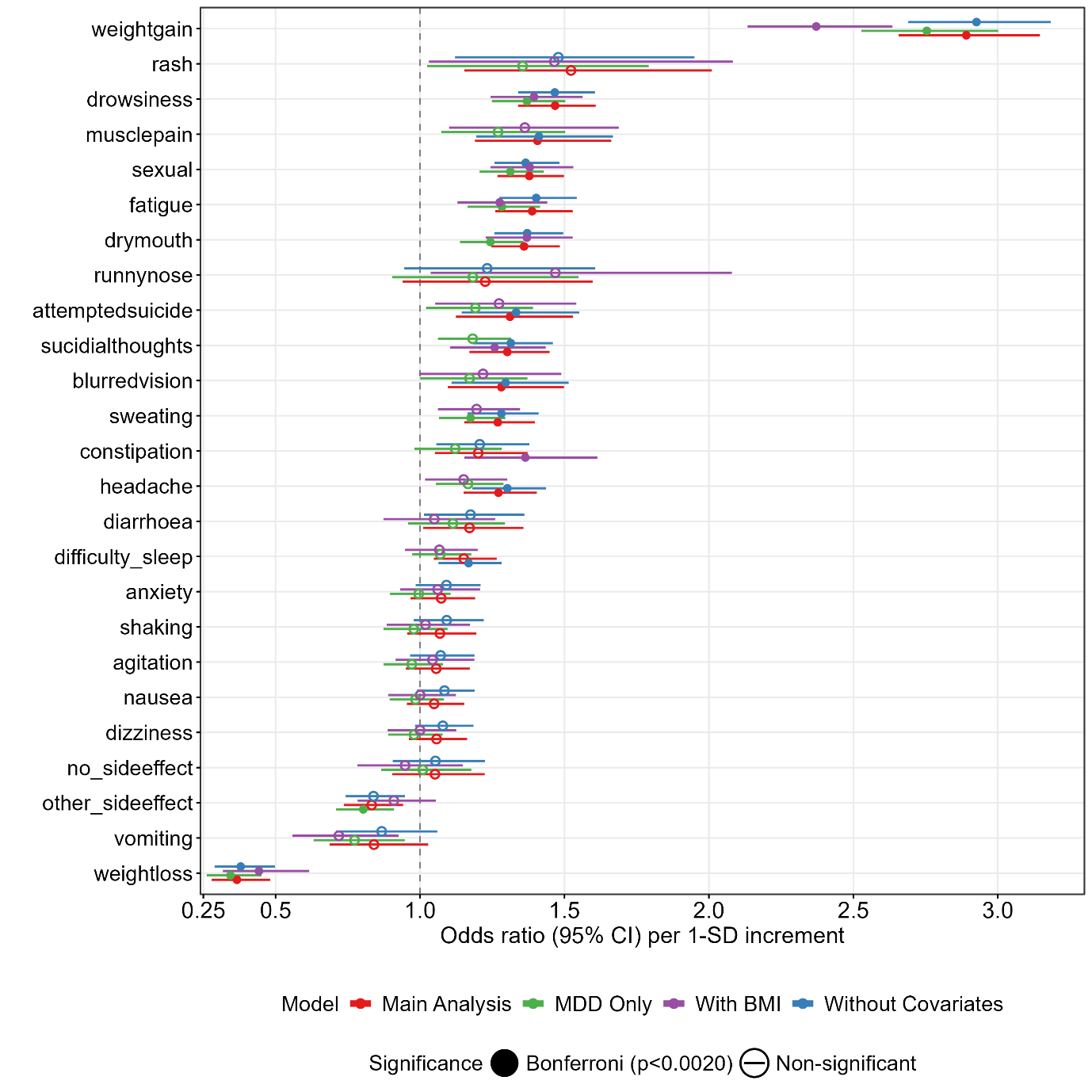

**Figure S5. Association between atypical depression and side effects of 10 antidepressant medications: main analysis and sensitivity analyses**

**Notes:** Sensitivity analyses included: (1) BMI-adjusted analysis (n=9,268) compared to main analysis controlling for age, sex and BMI (n=13,485); (2) MDD-confirmed cases only (n=12,010); (3) unadjusted model (n=13,495). Results show odds ratios from separate logistic regression models for each PGS with Bonferroni correction. Bars indicate 95% CI.

***Sensitivity analysis: MDD-confirmed cases***

Patterns were similar but effect sizes were attenuated in MDD-confirmed cases (Figure S1). ADHD-PGS (OR, 1.07; 95%CI, 1.02-1.12; p=0.0049) and MDD-PGS (OR, 1.08; 95%CI, 1.03-1.13; p=0.0009) remained significant, while Neuroticism-PGS (OR, 1.06; 95%CI, 1.01-1.11; p=0.015) did not survive Bonferroni correction, and BD-PGS are no longer significant. For physical health related PGS, all Bonferroni-significant associations remained for BMI, T2D, IR, CRP and HDL-C (Figure S2). For sleep/circadian traits, chronotype-PGS remained significant (OR, 0.94; 95%CI, 0.90-0.98; p=0.008).

Only SNRI remained significant after Bonferroni correction (OR, 0.88; 95%CI, 0.80-0.98; p=0.014), while SSRI and TCA associations were no longer significant. Significant side effects were reduced from twelve to eight: weight gain, drowsiness, sexual dysfunction, fatigue, dry mouth, sweating, and decreased weight loss. Associations with muscle pain, headache, sweating, and suicidal thoughts were no longer significant, while ‘other side effects’ became newly significant (Table S22; Figure S5).

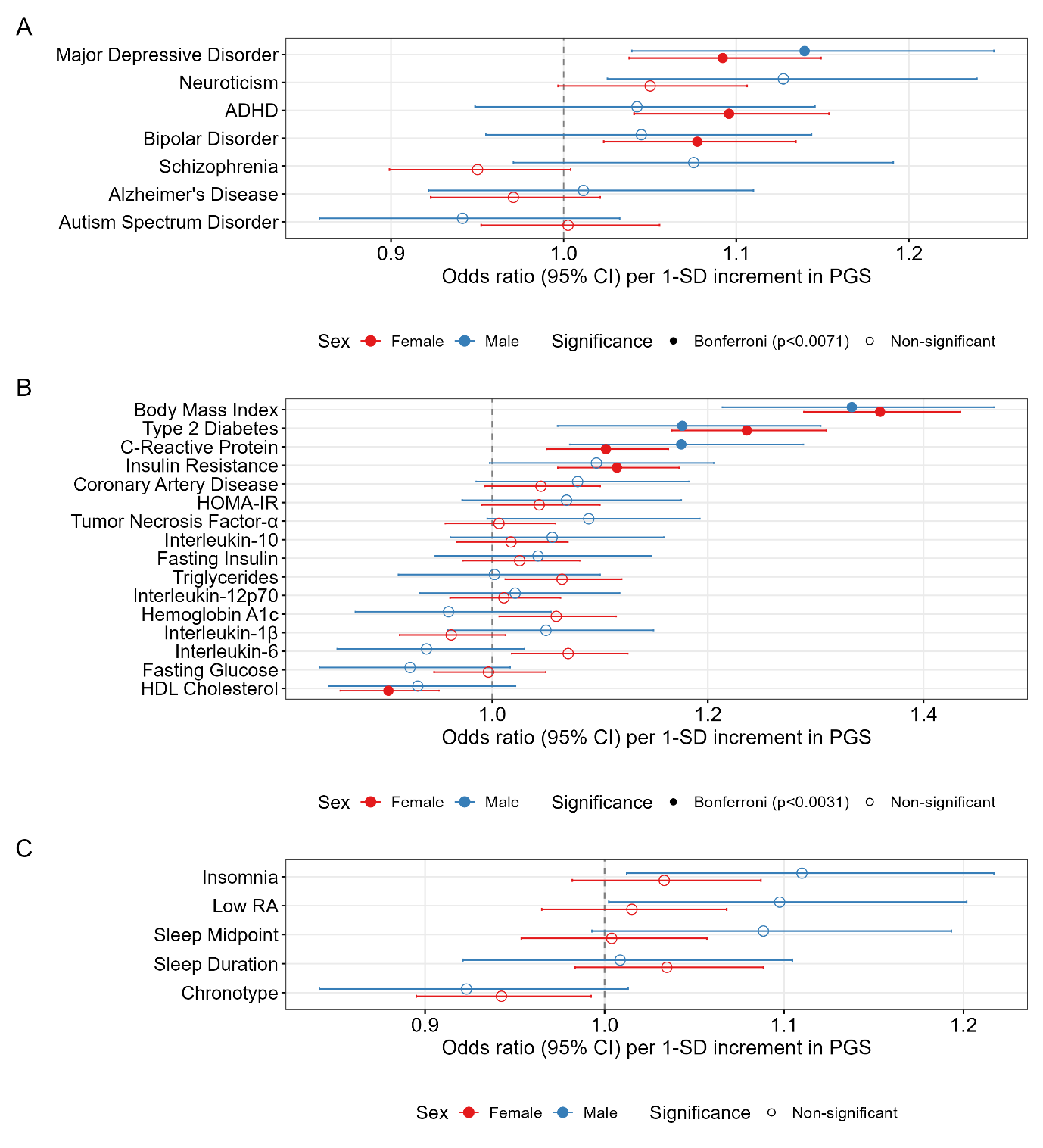

**Figure S6. Association between atypical depression and polygenic scores (PGS) by sex: (A) mental disorders; (B) metabolic and inflammatory traits; and (C) sleep and circadian traits (females: n=1,929 atypical, n=6,849 other depressive disorders; males: n=566 atypical, n=2,657 other depressive disorders).**

**Notes:** Results shown are each PGS with atypical depressive subtypes from separate regression models with covariates of age, sex, and the first 10 genetically-inferred ancestry PCs in logistic regression. Bars indicate 95% CI. Color-coding represents significance levels: Red: Bonferroni-corrected, and Gray: Non-significant.

**Abbreviations:** ADHD, Attention-Deficit Hyperactivity Disorder; HOMA-IR, Homeostasis model assessment of insulin resistance (derived from glucose and insulin); RA, Relative amplitude.

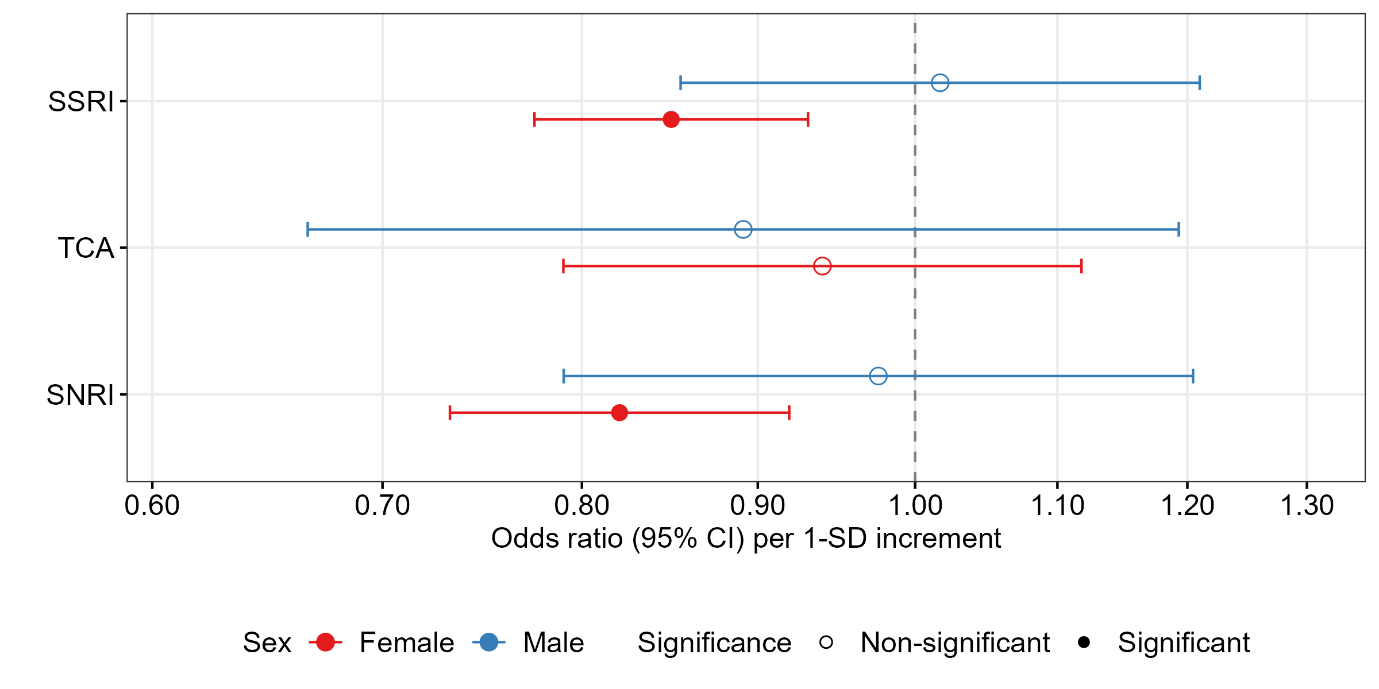

**Figure S7. Association between atypical depression and self-reported efficacy of antidepressants by sex**

**Notes**: Sex-stratified analysis controlling for age (males: SSRI n=2,814; SNRI n=1,769; TCA n=946; females: SSRI n=8,924; SNRI n=5,228; TCA n=2,402). Results show odds ratios from separate ordinal regression models for each PGS with Bonferroni correction. Bars indicate 95% CI.

**Abbreviations:** SSRI, Selective Serotonin Reuptake Inhibitors; SNRI, Serotonin-Norepinephrine Reuptake Inhibitors; TCA, Tricyclic Antidepressants.

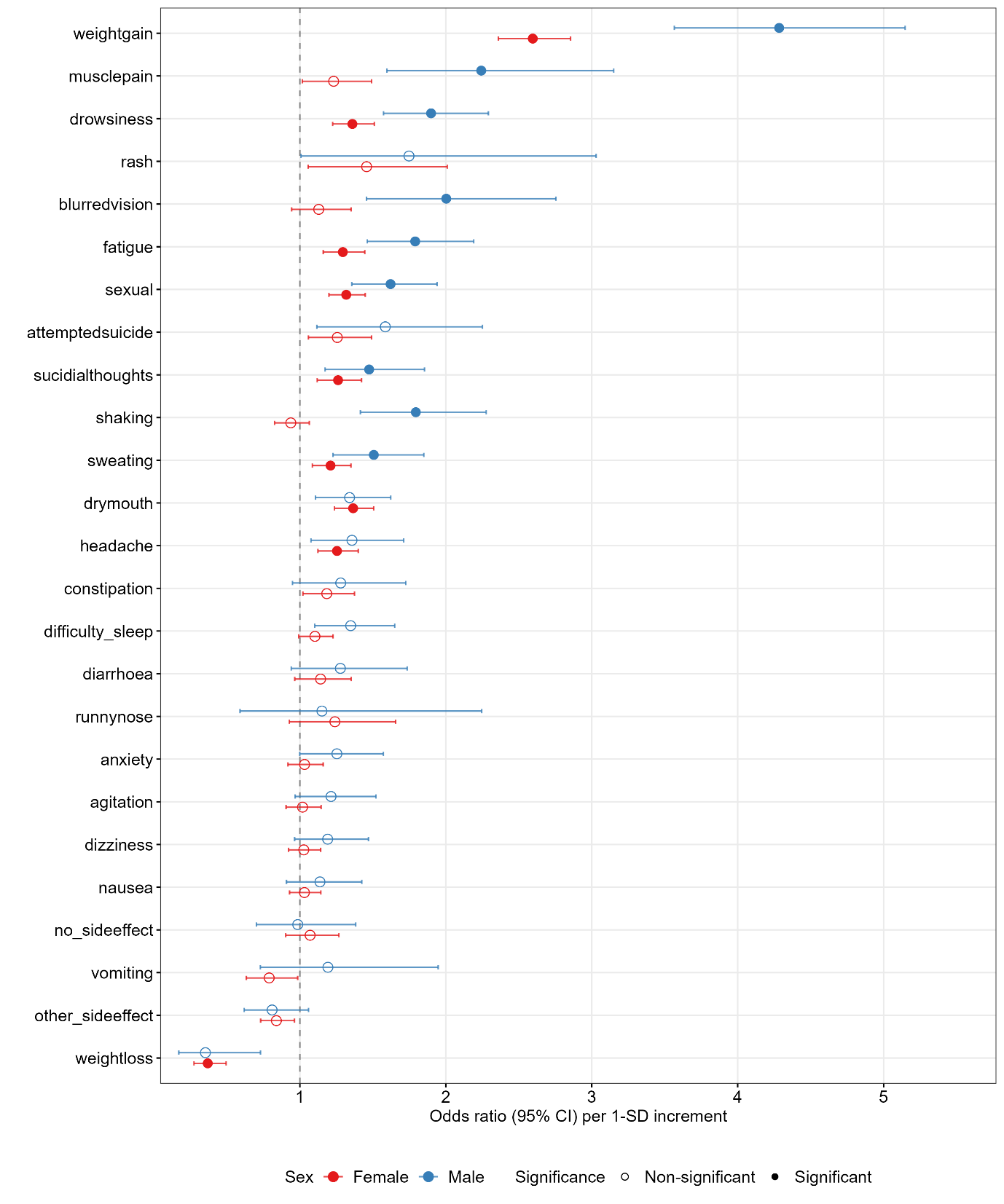

**Figure S8. Association between atypical depression and side effects of 10 antidepressant medications by sex**

**Notes**: Results shown are each antidepressant efficacy with atypical depressive subtypes from separate regression models with covariates of age in logistic regression. Sample sizes vary by specific side effect and sex. Bars indicate 95% CI.

**Sex-stratified analysis**

For PGS, we conducted additional sex-stratified analysis. Of the participants, 17.6% (n=566) were atypical cases within males and 22% (n=1,929) in females. In females, ADHD-PGS (OR, 1.10; 95%CI, 1.04-1.15; p=0.0005), MDD-PGS (OR, 1.09; 95%CI, 1.04-1.15; p=0.0007), and BD-PGS (OR, 1.08; 95%CI, 1.02-1.13; p=0.005) remained significant after Bonferroni correction. In males, only MDD-PGS (OR, 1.14; 95%CI, 1.04-1.25; p=0.005) survived correction, while ADHD-PGS, BD-PGS and Neuroticism-PGS were not significant. For physical health-related, both sexes remained significant associations with BMI-PGS (males: OR, 1.33; 95%CI, 1.21-1.47; p=2.22×10^-9^; females: OR, 1.36; 95%CI, 1.29-1.43; p=1.57×10^-29^), T2D-PGS (males: OR, 1.18; 95%CI, 1.06-1.30; p=0.002; females: OR, 1.24; 95%CI, 1.17-1.31; p=7.61×10^-13^) and CRP-PGS (males: OR, 1.18; 95%CI, 1.07-1.29; p=0.0006; females: OR, 1.11; 95%CI, 1.05-1.16; p=0.0001). HDL-C and IR-PGS were significant only in females. Chronotype-PGS showed marginal significance only in females (OR, 0.94; 95%CI, 0.90-0.99; p=0.025), while in males it was not significant (p=0.092). Overall, females showed more consistent PGS associations across mental health and metabolic domains, while males demonstrated fewer significant associations, suggesting potential sex-specific genetic architectures underlying atypical depression.

For antidepressant efficacy, no significant associations were found within males, while SSRI (OR, 0.85; 95%CI, 0.77-0.93; p=0.0005) and SNRI (OR, 0.82; 95%CI, 0.73-0.92; p=0.0006) efficacy remained significantly associated with atypical depression in females, likely reflecting the higher proportion of females in this sample (75%). For side effects, both sexes showed significant associations with multiple adverse effects, with considerable overlap. Common significant side effects in both sexes include drowsiness, fatigue, weight gain, reduced sexual dysfunction, and suicidal thoughts. Males additionally showed significant associations with sweating, shaking, muscle pain, and blurred vision, while females showed unique associations with dry mouth, headache, and weight loss. Males demonstrated 9 significant side effects compared to 9 in females, suggesting similar overall side effect burden despite some sex-specific patterns.

**References**

1.	American Psychiatric Association. Diagnostic and statistical manual of mental disorders: DSM-5: American psychiatric association Washington, DC; 2013.

2.	Altman EG, Hedeker D, Peterson JL, Davis JM. The Altman self-rating mania scale. Biological psychiatry. 1997;42(10):948-55.

3.	Konings M, Bak M, Hanssen M, Van Os J, Krabbendam L. Validity and reliability of the CAPE: a self‐report instrument for the measurement of psychotic experiences in the general population. Acta Psychiatrica Scandinavica. 2006;114(1):55-61.

4.	Van Spijker BA, Batterham PJ, Calear AL, Farrer L, Christensen H, Reynolds J, et al. The Suicidal Ideation Attributes Scale (SIDAS): Community‐based validation study of a new scale for the measurement of suicidal ideation. Suicide and Life‐Threatening Behavior. 2014;44(4):408-19.

5.	Kessler RC, Barker PR, Colpe LJ, Epstein JF, Gfroerer JC, Hiripi E, et al. Screening for serious mental illness in the general population. Archives of general psychiatry. 2003;60(2):184-9.

6.	Horne JA, Östberg O. A self-assessment questionnaire to determine morningness-eveningness in human circadian rhythms. International Journal of Chronobiology. 1976;4:97-110.

7.	Rosenthal NE, Sack DA, Gillin JC, Lewy AJ, Goodwin FK, Davenport Y, et al. Seasonal affective disorder. A description of the syndrome and preliminary findings with light therapy. Arch Gen Psychiatry. 1984;41(1):72-80.

8.	Demontis D, Walters GB, Athanasiadis G, Walters R, Therrien K, Nielsen TT, et al. Genome-wide analyses of ADHD identify 27 risk loci, refine the genetic architecture and implicate several cognitive domains. Nature Genetics. 2023;55(2):198-208.

9.	Bellenguez C, Küçükali F, Jansen IE, Kleineidam L, Moreno-Grau S, Amin N, et al. New insights into the genetic etiology of Alzheimer's disease and related dementias. Nat Genet. 2022;54(4):412-36.

10.	Grove J, Ripke S, Als TD, Mattheisen M, Walters RK, Won H, et al. Identification of common genetic risk variants for autism spectrum disorder. Nat Genet. 2019;51(3):431-44.

11.	O'Connell KS, Koromina M, van der Veen T, Boltz T, David FS, Yang JMK, et al. Genomics yields biological and phenotypic insights into bipolar disorder. Nature. 2025;639(8056):968-75.

12.	Adams MJ, Streit F, Meng X, Awasthi S, Adey BN, Choi KW, et al. Trans-ancestry genome-wide study of depression identifies 697 associations implicating cell types and pharmacotherapies. Cell. 2025;188(3):640-52.e9.

13.	Nagel M, Jansen PR, Stringer S, Watanabe K, de Leeuw CA, Bryois J, et al. Meta-analysis of genome-wide association studies for neuroticism in 449,484 individuals identifies novel genetic loci and pathways. Nature Genetics. 2018;50(7):920-7.

14.	Trubetskoy V, Pardiñas AF, Qi T, Panagiotaropoulou G, Awasthi S, Bigdeli TB, et al. Mapping genomic loci implicates genes and synaptic biology in schizophrenia. Nature. 2022;604(7906):502-8.

15.	Yengo L, Sidorenko J, Kemper KE, Zheng Z, Wood AR, Weedon MN, et al. Meta-analysis of genome-wide association studies for height and body mass index in ∼700000 individuals of European ancestry. Human Molecular Genetics. 2018;27(20):3641-9.

16.	Aragam KG, Jiang T, Goel A, Kanoni S, Wolford BN, Atri DS, et al. Discovery and systematic characterization of risk variants and genes for coronary artery disease in over a million participants. Nat Genet. 2022;54(12):1803-15.

17.	Dupuis J, Langenberg C, Prokopenko I, Saxena R, Soranzo N, Jackson AU, et al. New genetic loci implicated in fasting glucose homeostasis and their impact on type 2 diabetes risk. Nat Genet. 2010;42(2):105-16.

18.	Lagou V, Mägi R, Hottenga J-J, Grallert H, Perry JRB, Bouatia-Naji N, et al. Sex-dimorphic genetic effects and novel loci for fasting glucose and insulin variability. Nature Communications. 2021;12(1):24.

19.	Sinnott-Armstrong N, Tanigawa Y, Amar D, Mars N, Benner C, Aguirre M, et al. Genetics of 35 blood and urine biomarkers in the UK Biobank. Nature Genetics. 2021;53(2):185-94.

20.	Oliveri A, Rebernick RJ, Kuppa A, Pant A, Chen Y, Du X, et al. Comprehensive genetic study of the insulin resistance marker TG:HDL-C in the UK Biobank. Nat Genet. 2024;56(2):212-21.

21.	Suzuki K, Hatzikotoulas K, Southam L, Taylor HJ, Yin X, Lorenz KM, et al. Genetic drivers of heterogeneity in type 2 diabetes pathophysiology. Nature. 2024;627(8003):347-57.

22.	Said S, Pazoki R, Karhunen V, Võsa U, Ligthart S, Bodinier B, et al. Genetic analysis of over half a million people characterises C-reactive protein loci. Nature Communications. 2022;13(1):2198.

23.	Sun BB, Chiou J, Traylor M, Benner C, Hsu Y-H, Richardson TG, et al. Plasma proteomic associations with genetics and health in the UK Biobank. Nature. 2023;622(7982):329-38.

24.	Jones SE, Lane JM, Wood AR, van Hees VT, Tyrrell J, Beaumont RN, et al. Genome-wide association analyses of chronotype in 697,828 individuals provides insights into circadian rhythms. Nat Commun. 2019;10(1):343.

25.	Ferguson A, Lyall LM, Ward J, Strawbridge RJ, Cullen B, Graham N, et al. Genome-Wide Association Study of Circadian Rhythmicity in 71,500 UK Biobank Participants and Polygenic Association with Mood Instability. EBioMedicine. 2018;35:279-87.

26.	Jansen PR, Watanabe K, Stringer S, Skene N, Bryois J, Hammerschlag AR, et al. Genome-wide analysis of insomnia in 1,331,010 individuals identifies new risk loci and functional pathways. Nat Genet. 2019;51(3):394-403.
